# Estimation and interpretation of vaccine efficacy in COVID-19 randomized clinical trials

**DOI:** 10.1101/2022.02.02.22270317

**Authors:** Hege Michiels, An Vandebosch, Stijn Vansteelandt

## Abstract

An exceptional effort by the scientific community has led to the development of multiple vaccines against COVID-19. Efficacy estimates for these vaccines have been widely communicated to the general public, but are nonetheless challenging to compare because they are based on phase 3 trials that differ in study design, definition of vaccine efficacy and in the handling of cases arising shortly after vaccination. In this work, we investigate the impact of these choices on vaccine efficacy estimates, both theoretically and by re-analyzing the Janssen and Pfizer COVID-19 trial data under a uniform protocol. We moreover study the causal interpretation that can be assigned to per-protocol analyses typically performed in vaccine trials. Finally, we propose alternative estimands to measure the intrinsic vaccine efficacy in settings with delayed immune response.

## 1 Introduction

The SARS-CoV-2 pandemic presents an extraordinary challenge to global health. In an attempt to curb the spread of disease and to control the pandemic, multiple COVID-19 vaccine candidates have been developed [35]. For example, FDA issued emergency use authorizations for two mRNA vaccines, developed by Pfizer and Moderna, in December 2020 [11, 12] and an adenovector vaccine, developed by Janssen, in February 2021 [13], based on safety and efficacy data from phase 3 randomized double-blind, placebo-controlled trials. The demonstration of vaccine efficacy and safety in phase 3 trials is essential for authorization and to inform policy makers about potential uses of vaccines [33]. Vaccine efficacy is defined as the relative reduction of the risk of disease for vaccinated participants compared to controls [14]. The efficacy estimates from the aforementioned trials far exceeded the FDA and WHO thresholds of an observed 50% reduction of symptomatic disease with a lower limit above 30% for the confidence interval [10, 34]. However, different risk measures, such as cumulative incidence, hazard or incidence rate have been used [33], making the obtained efficacy estimates possibly difficult to compare.

Although the intention-to-treat principle is central to clinical trial research, vaccine efficacy trials usually conduct a per-protocol approach where the population is restricted to eligible, fully compliant participants receiving all doses [18, 33]. These analyses moreover include a delay, typically starting after the last dose of the vaccine plus the maximum incubation period, to allow the immune response to develop and to account for the time between infection and symptom onset [5]. The delay between vaccination and the development of a robust immune response is referred to as the vaccine ramp-up time [6]. Per-protocol analyses aim to estimate the intrinsic efficacy of the vaccine, but are vulnerable to post-randomization biases [5].

In this article, we mainly focus on trials of COVID-19 candidate vaccines although the methods are generalizable to any vaccine trial. In particular, we discuss studies of AstraZeneca [9], Janssen [13], Moderna [11] and Pfizer [12]. Table 1 describes the design and efficacy results of COVID-19 trials conducted by these companies. These trials have variable dosing regimens, endpoints, risk measures, ramp-up periods and follow-up times. The differences in study length make it particularly difficult to compare vaccine efficacy estimates as the virus mutates [23]. In addition, these trials consider different populations, vaccine efficacy estimators and disagree in the way intercurrent events are handled [1, 24, 28, 32]. For example, the primary analysis of the Janssen trial investigated the efficacy of the experimental vaccine against confirmed moderate to severe/critical COVID-19 [28], while the other companies included all confirmed cases [1, 24, 32]. Moreover, the Moderna study [1] was conducted at different centers in the United States, while the other studies took place on sites across different continents. Consequently, the distribution of the ethnic groups of trial participants is also different (figure 1 in Appendix A). All trials had a case-driven study design, requiring a particular number of cases to trigger the primary analysis [9, 11, 12, 13].

**Table 1:** Design and reported results of COVID-19 vaccine trials. Information is based on the following publications: AstraZeneca: pooled analysis of ISRCTN89951424 and ClinicalTrials.gov NCT04400838 [32] Janssen: ClinicalTrials.gov NCT04505722 [28] Moderna: ClinicalTrials.gov NCT04470427 [1] Pfizer: ClinicalTrials.gov NCT04368728 [24]

|  | **AstraZeneca** | **Janssen** | **Moderna** | **Pfizer** |
| --- | --- | --- | --- | --- |
| **Dosing regimen** | Two doses of ChAdOx1 nCoV-19 or placebo<br>28 days apart | Single dose of Ad26.COV2.S or placebo | Two intramuscular injections of mRNA-1273 or placebo<br>28 days apart | Two doses of BNT162b2 or placebo<br>21 days apart |
| **Endpoint** | Virologically confirmed, symptomatic COVID-19 | Confirmed moderate to severe/critical COVID-19 | Symptomatic COVID-19 | Confirmed COVID-19 |
| **Risk measure** | Incidence rate (Poisson model) | Incidence rate (Poisson model) | Hazard rate (Cox model) | Incidence rate (Bayesian beta-binomial model) |
| **Ramp-up period** | Infections before day 15 after second dose were removed | Infections before day 14 after vaccination were removed | Infections before day 14 after second dose were censored | Infections before day 7 after second dose were removed |
| **Follow-up** | Average: 43 days | Median: 58 days<br>Range: [1; 124] | Median: 63 days<br>Range: [0; 97]<br>(after second dose) | Average: 46 days<br>(starting from 7 days after second dose) |
| **Vaccine efficacy (95% CI)** | 70.4% ([54.8; 80.6]) | 66.9% ([59.0; 73.4]) | 94.1% ([89.3; 96.8]) | 95.0% ([90.3; 97.6]) |

**Figure 1:**
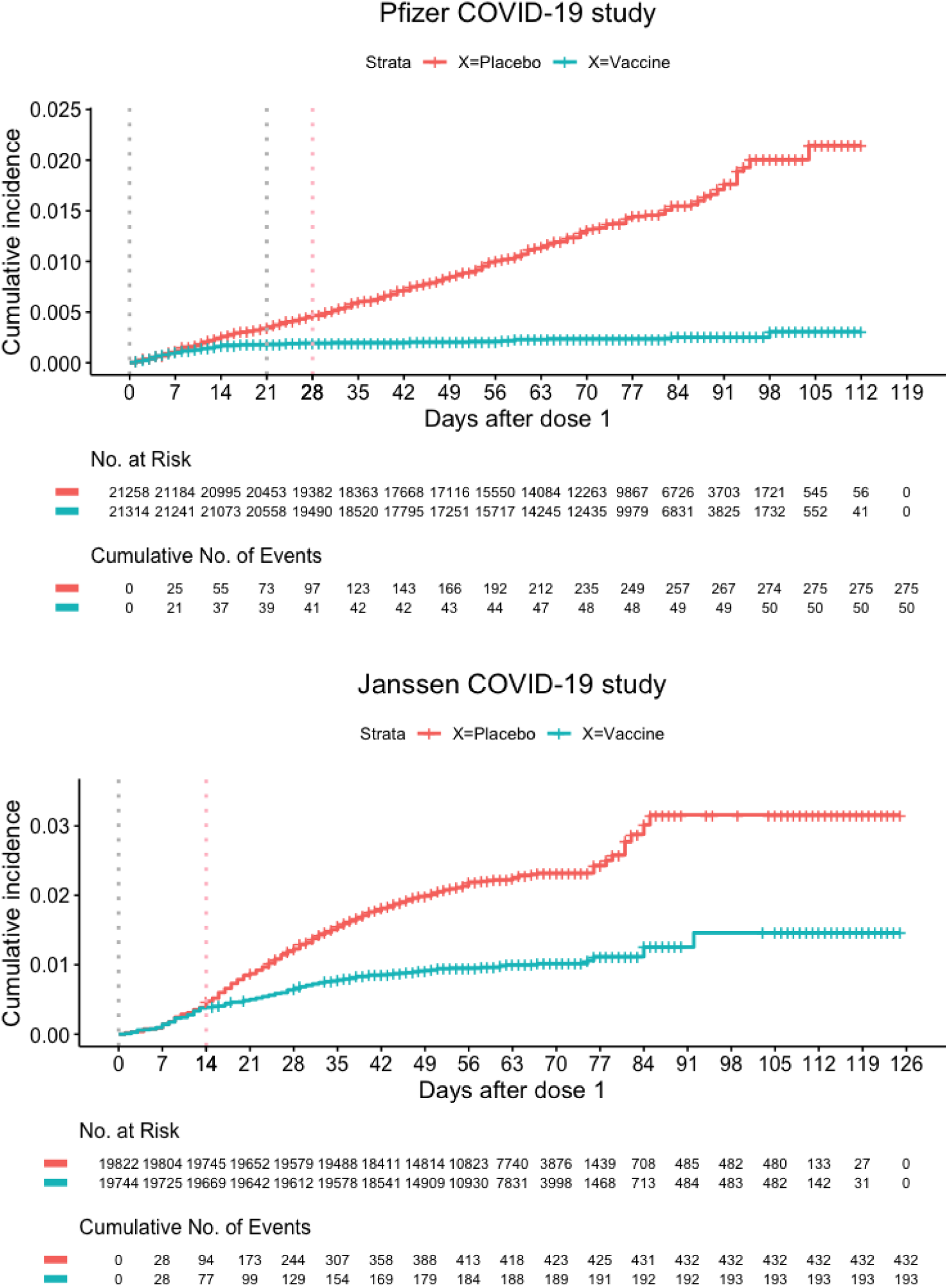
Recreated data similar to the COVID-19 vaccine trials conducted by Pfizer [12] and Janssen [13]. The grey vertical lines indicate the visits at which a dose of the vaccine is given and the pink line indicates the end of the ramp-up period, as specified in the study protocol.

Rapaka et al. [25] compared different phase 3 COVID-19 vaccine trials and concluded that comparisons of vaccine efficacy estimates must be made with careful consideration because of the differences in study design, study population and characteristics of circulating virus variants. In this paper, we investigate the implications of different choices made in the statistical analyses (rather than the study design or study population). In particular, we review the impact of the choice of the risk measure, investigate the change in vaccine efficacy over time and examine how cases arising shortly after vaccination are handled, both theoretically and by re-analyzing the Janssen and Pfizer COVID-19 data. In addition, we develop insight into the causal interpretation of different definitions of vaccine efficacy and propose alternative estimands to measure vaccine efficacy in settings with delayed immune response.

## 2 Vaccine efficacy

The primary efficacy endpoint in phase 3 vaccine trials is often defined with respect to clinical disease with laboratory confirmation, since the goal of vaccination is typically to prevent disease and not necessarily to prevent infection [18, 33]. Vaccine efficacy (VE) typically has the form VE = 1-RR, with RR a measure of relative risk of disease in vaccinated subjects compared to placebo subjects. This measure of vaccine efficacy takes values in the interval ] − ∞, 1], with 1 indicating complete protection by the vaccine, 0 expressing no effect, and a negative value representing an increase in risk of disease due to vaccination [18]. As risk measure the cumulative incidence, incidence or hazard rate is typically used [15].

When using the cumulative incidence, the vaccine efficacy represents the relative reduction in risk of developing disease during the duration of the trial attributable to vaccination [18]. It has been argued that this definition of vaccine efficacy is especially appropriate if the vaccine has an ‘all-or-nothing’ mode of action, meaning that vaccination renders a part of the population completely immune, while offering no protection for the remainder [29, 18]. This is because it can be interpreted as a number of cases that can be avoided by vaccination, or the probability that vaccination prevents infection before the considered time, for individuals who would be infected before that time if not vaccinated (Appendix B.1). This interpretation is justified under the assumption that vaccination can never shorten the infection times. Without this assumption, the vaccine efficacy definition using cumulative incidence can still be interpreted as a lower bound for this probability (Appendix B.1).

The interpretation of vaccine efficacy estimates using the incidence or hazard rate as risk measure is not straightforward. In particular, these effects cannot easily be transferred to a proportion of cases that can be avoided by vaccination. However, these measures of vaccine efficacy are arguably useful if the vaccine is ‘leaky’, meaning that vaccination reduces the hazard of the disease by a multiplicative factor for all vaccinated subjects [29, 18]. The mode of action of a vaccine is usually unknown, but for rare diseases with constant incidence rate, all these risk measures lead to approximately the same vaccine efficacy estimands [18]. In Appendix B.2, we illustrate this by approximating the survival functions using Taylor expansions. The variance of the vaccine efficacy defined using cumulative incidence, usually estimated by the Kaplan-Meier estimator, is expected to be larger than when using the hazard or incidence rate, because of the (semi-)parametric nature of the Cox and Poisson model.

## 3 Delayed immunization

Preventive vaccine trials typically employ a per-protocol (PP) approach wherein only fully compliant patients are included [18, 33]. In addition, cases are often defined as patients who became ill after a fixed time lag beyond randomization in order to take into account the incubation period and to allow the vaccinee to develop a protective immune response [18, 33]. For example, the per-protocol analysis in the Janssen COVID-19 trial was restricted to cases occurring at least 14 and 28 days after vaccination respectively [13]. Infections observed before these days were removed from the analysis set. In this section, we investigate the effect of taking into account this additional vaccine ramp-up time for immunity to develop.

### 3.1 Per-protocol effects in COVID-19 trials

The aim of per-protocol analyses is to obtain insight into the intrinsic efficacy of the vaccine, i.e. the relative reduction in infections due to vaccination in fully compliant subjects, after completion of the prescribed vaccination regimen and achieving adequate immune response [17, 18]. However, this approach can negatively impact power relative to including all cases observed since baseline, especially in settings with low disease incidence [6]. It is moreover vulnerable to selection bias as per-protocol analyses entail comparison of subgroups selected post randomization [18].

In the Janssen trial, the cumulative incidence in the full analysis set was similar in both the vaccine and placebo arm until around day 14, after which the curves diverged, with more cases accumulating in the placebo group than the vaccine group (figure 1). Therefore, we expect the possible selection bias in the per-protocol analysis to be more severe when cases observed before day 28 are removed compared to when cases before day 14 are removed. Selection bias may occur if cases develop during the ramp-up period and vaccinated and placebo groups are no longer at comparable risk of infection after this period [17]. In the Janssen trial, approximately 18% of the placebo cases and 39% of the vaccine cases were observed during the first 14 days (figure 1), even though it is expected that only a small proportion of the total cases occurs shortly after randomization [17]. If the vaccine has no effect on infections and there are no side effects during the ramp-up period, it is expected that removing cases observed during this period does not introduce selection bias, as these cases are then likely comparable across arms. One then obtains the vaccine effect for a subgroup of the study population so that the per-protocol effect may nevertheless differ from the intention-to-treat (ITT) effect that takes into account all cases after randomization (Appendix C.1).

Other pharmaceutical companies also included some time lag beyond completion of the last vaccination dose to allow for optimal immunity (table 1). In the Pfizer trial, the cumulative incidence in the full analysis set was again similar in both arms until approximately 14 days after randomization, at which time point the survival curves diverged (figure 1). However, a ramp-up period of 28 days (after randomization) was specified, which resulted in the removal of approximately 34% of the placebo and 82% of the vaccine cases (figure 1). For the AstraZeneca and Moderna trials, it also appears that the vaccine already had a clear effect on infection before the end of the specified ramp-up period (Figure 7 in [9] and Figure 2 in [11]).

**Figure 2:**
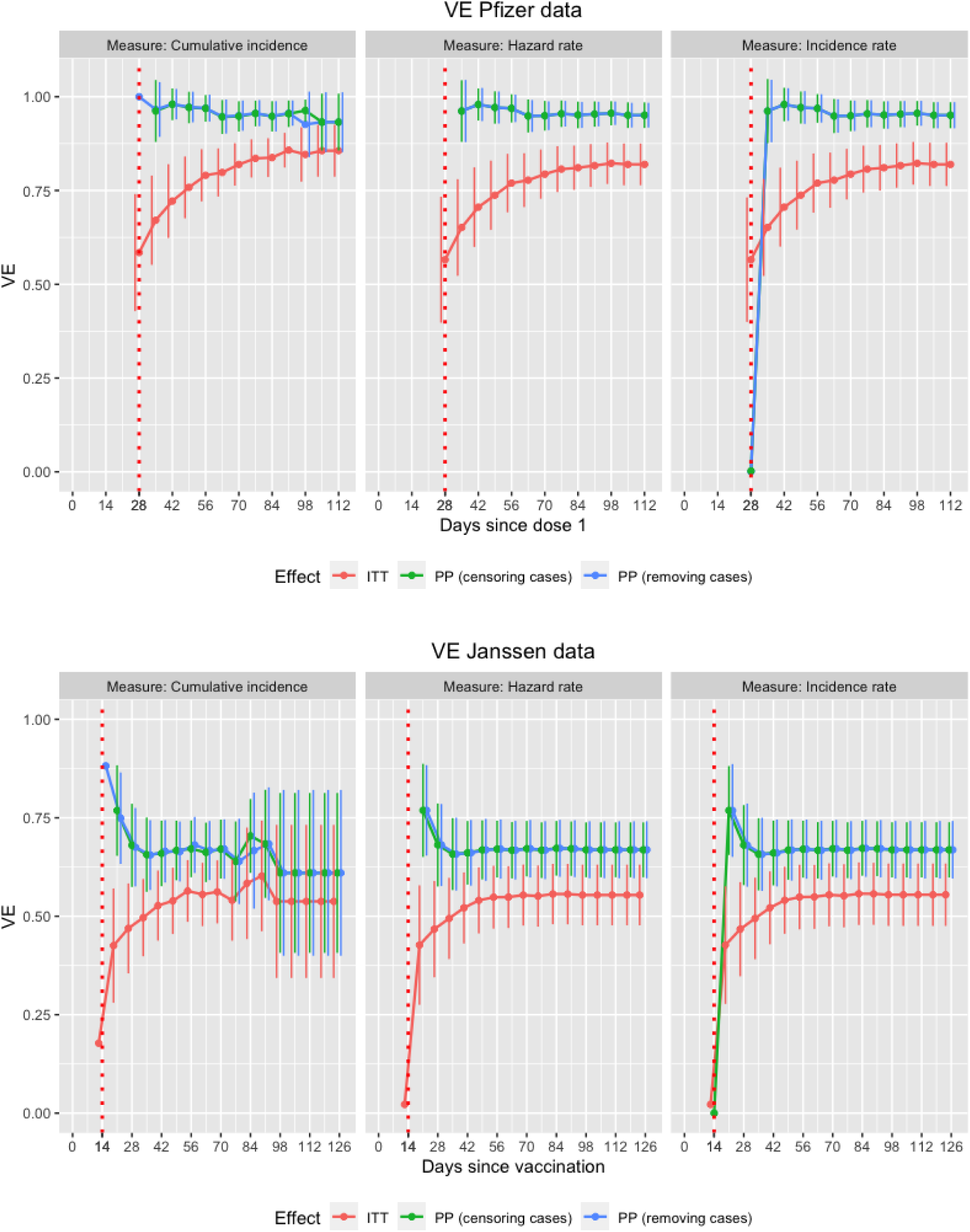
Vaccine efficacy estimates ± 2SE for the Pfizer and Janssen dataset are shown over time, both for the cumulative incidence, the hazard and incidence rate as risk measure. Every point represents the VE estimate that would be obtained if the trial were stopped at the corresponding visit and all information up till that visit was used.

In the Moderna trial, vaccine efficacy was estimated using a hazard ratio that was obtained by fitting a stratified Cox proportional hazards model [4], where patients who got infected before day 14 after the second dose were censored [11]. This approach is also subject to a possible selection bias, since the implicit assumption of non-informative censoring is violated as cases are censored based on their infection time. None of the aforementioned pharmaceutical companies reported a rationale for the choice of the length of the ramp-up period in the protocol [9, 13, 11, 12], even though it is recommended by the WHO [33].

The WHO [33] argued that, in general, the ITT estimate will tend to be diluted com-pared to the PP vaccine efficacy estimates, since individuals typically fail to comply with the protocol for reasons related to the vaccine itself [5]. In addition, including cases that arise during the ramp-up time will typically lead to smaller vaccine efficacy estimates since the vaccine is not yet fully effective during this period. Since ITT vaccine efficacy estimates provide information about the effectiveness of a public health strategy using the vaccine, and because they reflect the speed at which the vaccine becomes protective, they may be more meaningful than per-protocol effects to compare vaccines with different dosing regimens or ramp-up periods [33]. To summarize, it has been recommended to report both vaccine efficacy estimates [17].

### 3.2 Hypothetical vaccine efficacy estimands

In this section, we propose two new estimands that can be used to measure vaccine efficacy in settings with delayed immune response and give insight into the intrinsic effect of the vaccine after achieving adequate immune response. In addition, we discuss new estimators for these estimands and clarify under what assumptions they can be approximated by standard per-protocol estimators.

#### 3.2.1 Vaccine efficacy if infections during ramp-up can be prevented

First, we consider the vaccine efficacy that would have been observed if cases during the ramp-up period could have been avoided. This is an example of a hypothetical estimand [19] and might be of particular interest since it is an effect that can be realized in practice for several viruses. For example, influenza vaccines cause antibodies to develop in the body about two weeks after vaccination [3]; influenza infections during this period can be avoided by vaccinating people early enough, i.e. at least two weeks before flu season begins. Further, vaccines against diseases endemic to certain countries, e.g. Malaria in Africa [2], are given early enough before traveling, allowing the immune response to be developed before arriving in these countries. In the COVID-19 setting, cases occurring shortly after vaccination can be avoided by quarantine. In general, we consider a setting where cases during the ramp-up period can be avoided.

It is not immediately clear how this effect can be identified without relying on strong assumptions. In particular, the observed infection hazard ratios after the ramp-up period cannot simply be used as substitution for the infection hazard ratio in the hypothetical setting because the populations not at risk for infections at a given time are likely not exchangeable between the observed and hypothetical setting. In Appendix C.2, we show that the per-protocol estimator, which removes cases observed during the ramp-up time, is unbiased for this effect only under strong assumptions. In particular, one must assume that patients who are infected during the ramp-up time would have comparable infection times as patients who were not infected during this period, if infections during this period could have been avoided. This is likely implausible as early cases may well be selective.

#### 3.2.2 Vaccine efficacy if ramp-up period can be eliminated

Next, we consider the vaccine efficacy, if the vaccine would immediately induce an immune response; i.e. if there was no ramp-up period. This estimand provides insight into the intrinsic effect of the vaccine because it represents the effect once subjects are fully immunized.

In Appendix C.3, we propose an estimator for this hypothetical VE which relies on a Structural Distribution Model (SDM) for identification [26, 31]. This model maps percentiles of the distribution of infection times under placebo into percentiles of the distribution of infection times under vaccine. In particular, if the vaccine would work immediately, we would impose that vaccination multiplies the quantiles of the infection distribution by a factor exp(*ψ*), for a scalar *ψ*. However, we assume that the vaccine effect is limited during the ramp-up time, and therefore, the quantiles are multiplied by a (possibly) smaller factor exp(*ρψ*) with *ρ* ∈ [0, 1]. In particular, let *α* denote the length of the ramp-up time, *S*(*t*|*X* = 0) the survival function in the placebo arm and *S*_SDM_(*t*|*X* = 1; *α, ρ, ψ*) the survival function in the vaccine arm. If we assume that the vaccine is fully effective after the ramp-up period, this leads to model

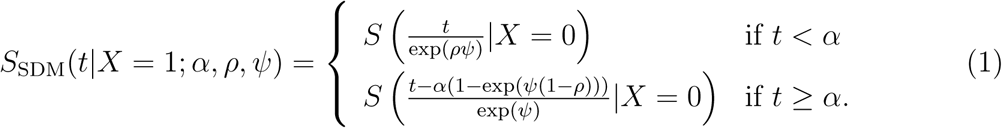

In this model, *ψ* represents the vaccine effect, with higher values indicating higher efficacy. Parameter *ρ* indicates how much weaker the vaccine effect is during the ramp-up time than after. The choice *ρ* = 0 expresses no vaccine effect during the ramp-up period, while *ρ* = 1 indicates full vaccine effect from baseline. Model (1) allows to impose that the vaccine has a different effect during and after the ramp-up time. The parameters can be estimated by comparing the mapped survival function (1) to the observed survival function in the vaccine arm. The hypothetical vaccine efficacy can then be estimated by setting the ramp-up period to 0 days in model (1). Details about this model and estimation can be found in Appendix C.3.

## 4 Data analysis

In this section, we compare the discussed vaccine efficacy estimands and risk measures by performing data analyses on data similar to the Janssen and Pfizer COVID-19 trials.

### 4.1 Data

The data of the Janssen and Pfizer COVID-19 trials were recreated, based on the published Kaplan-Meier curves (Figure 1 in [13] and Figure 2 in [12]), as described in Appendix C.4.1. Figure 1 visualizes the obtained Kaplan-Meier curves, which agree very well with the published curves. The R-code used to create these datasets is provided in Appendix D.1.

### 4.2 Methods

For both trials, the ITT and PP vaccine efficacy effects were estimated every week for the entire study duration, using the cumulative incidence, the hazard and incidence rate as risk measures. ITT effects included all cases observed since randomization, while for the PP effects cases observed during the ramp-up time were removed or censored. Different lengths of ramp-up times (*α* = 7, 14, 28 and 35 days) were investigated for both trials. Hazard ratios were estimated by fitting a Cox proportional hazards model [4] and cumulative incidences were obtained by estimating Kaplan-Meier curves [21]. Incidence rates were acquired by fitting a Poisson model [22] with the logarithm of the observation time as offset to account for follow-up time. All estimators assume censoring to be non-informative within each treatment arm. The hypothetical estimand ‘if the ramp-up period could be eliminated’ (section 3.2.2) was also estimated every week, using the cumulative incidence as risk measure, as described in Appendix C.4.3. R-code for these estimators is provided in Appendix D.2. Standard errors (SE) were obtained using 1000 non-parametric bootstrap replications [8].

### 4.3 Results

Table 2 shows the obtained vaccine efficacy estimates and SEs for the Pfizer and Janssen COVID-19 vaccine trials, using the ramp-up period as specified in the study protocols. For both trials, the ITT effect estimates are approximately 10% smaller than the PP effects. This is in contrast with the results of Horne et al. [17] who generally observed little difference between ITT and PP effects. However, these authors also found trials reported in the last 20 years where efficacy estimates under the two approaches gave discordant results.

**Table 2:** Results of the data analysis performed on the Pfizer and Janssen dataset. Pfizer: vaccine efficacy is measured at day 112 and the length of the ramp-up period is *α* = 28 days. Janssen: vaccine efficacy is measured at day 125 and the length of the ramp-up period is *α* = 14 days.

| Effect | Risk measure | Vaccine efficacy estimate (SE) |  |
| --- | --- | --- | --- |
|  |  | Pfizer | Janssen |
| Intention-to-treat | Hazard rate | 0.82 (0.03) | 0.55 (0.04) |
|  | Cumulative incidence | 0.86 (0.03) | 0.54 (0.10) |
|  | Incidence rate | 0.82 (0.03) | 0.55 (0.04) |
| Per-protocol<br>(removing cases before $\alpha$) | Hazard rate | 0.95 (0.02) | 0.67 (0.04) |
|  | Cumulative incidence | 0.93 (0.04) | 0.61 (0.11) |
|  | Incidence rate | 0.95 (0.02) | 0.67 (0.04) |
| Per-protocol<br>(censoring cases before $\alpha$) | Hazard rate | 0.95 (0.02) | 0.67 (0.03) |
|  | Cumulative incidence | 0.93 (0.04) | 0.61 (0.10) |
|  | Incidence rate | 0.95 (0.02) | 0.67 (0.04) |
| VE if ramp-up can be eliminated | Cumulative incidence | 0.93 (0.04) | 0.66 (0.13) |

In addition, PP effects where cases observed during the ramp-up period are removed versus censored coincide in our results. However, the vaccine efficacy estimates differ by the risk measure used, even though that was not expected because of the small incidence rates [18]. Moreover, for most effects, the obtained standard errors were two to three times larger when using the cumulative incidence compared to the hazard or incidence rate. This can be attributed to the (semi-)parametric nature of the Cox and Poisson model, but comes at the risk of bias when this model is not correct. Figure 2 shows that the effect estimates converge over time. In the Janssen trial, an increase in VE was noticed 84-91 days after vaccination when using the cumulative incidence as risk measure. This is probably because many patients are censored during that period (figure 1) and is not observed when using the other risk measures.

Tables 1 and 2 and figures 10 and 11 in Appendix C.4.2 show the results when using other lengths of the ramp-up period. From these results, it turns out that choosing too short a period is more problematic than too long a period. In particular, when *α* is set to 7 days in the Janssen trial and the Pfizer trial, diluted vaccine efficacy estimates are obtained compared to the original PP effects. In both trials, the PP effects converge to approximately the same limit when specifying a ramp-up period of 14, 28 or 35 days. However, in the Janssen trial, the obtained SEs are somewhat larger when more cases are removed/censored.

Table 2 also shows the estimates for the hypothetical estimand ‘if there was no ramp-up time’. Although different estimators were used (Appendix C.4.3), we only show results for the one-parameter model method with the ramp-up period specified as in the study protocol. The obtained effect estimates are very close to the PP estimates with only a slightly larger SE (table 2). For the Pfizer trial, the Structural Distribution Model fitted the observed data very well (figures 13 - 15 in Appendix C.4.3), but less so for the Janssen study (figures 17 - 18 in Appendix C.4.3).

## 5 Discussion

In this paper, we compared the primary estimand and estimator, based on the per-protocol principle of different phase 3 trials with four investigational vaccines, with a common objective to evaluate efficacy and safety in preventing COVID-19. In particular, the different vaccine efficacy estimands have been investigated and discussed. Using cumulative incidence as risk measure is the most interesting in terms of interpretation, since the corresponding vaccine efficacy represents a number of cases that can be avoided by vaccination. The hazard and incidence rate do not lead to a straightforward interpretation of vaccine efficacy, but show more stability when estimated (semi-)parametrically and result in approximately the same vaccine efficacy estimates in most settings with rare diseases. However, in the data analyses performed on the recreated Janssen and Pfizer COVID-19 trials, we obtained differences up to 6 percent points, depending on the chosen risk measure.

We also observed differences of approximately 10 percent points between intention-to-treat analyses, taking into account all cases since baseline, and per-protocol analyses, removing or censoring cases occurring shortly after vaccination, while other authors generally found little difference [17]. Since per-protocol analyses are subject to possible selection bias [5] and are not aligned with a relevant estimand [19], we have proposed two hypothetical estimands that give insight into the intrinsic effect of the vaccine in settings with delayed immune response. The first estimand considers the vaccine efficacy that would have been observed if cases during the ramp-up could be prevented. We argued that strong assumptions are needed for the per-protocol analysis to unbiasedly estimate this effect. The second estimand considers the vaccine efficacy if the ramp-up period can be eliminated. We developed a novel estimator for this estimand using a Structural Distribution Model [26, 31]. This proposal relies on modeling assumptions, which can partially be checked by comparing the modeled vaccine survival curve to the observed curve. In contrast to the principal stratification estimand, the proposed estimand targets an effect for the entire trial population. In addition, our proposal does not rely on monotonicity or principal ignorability assumptions that are typically needed to estimate principal stratification effects [7].

To conclude, the vaccine efficacy estimates obtained in the different COVID-19 trials are quite comparable (except for the differences in study design). These per-protocol vaccine efficacy estimates can, in this setting, be interpreted as the number of cases that can be avoided by vaccination if the vaccine would immediately induce an immune response The use of naive per-protocol effects may be problematic in other settings, in which case we recommend estimation of the proposed estimand.

The conclusions drawn in this article are not only useful in the COVID-19 trial setting, but can also be applied to other vaccine efficacy trials.

## Data Availability

The data are not publicly available due to confidentiality restrictions. The simulated data that support the findings of this
study are available on request from the corresponding author.

## Acknowledgments

This work was supported by VLAIO (Flemish Innovation and Entrepreneurship) [Baekeland grant agreement HBC.2019.2155].

## Conflict of interest

The authors declare no potential conflict of interests.

## Notes

### Competing Interest Statement

The authors have declared no competing interest.

