## Appendix for "Estimation and interpretation of vaccine efficacy in COVID-19 randomized clinical trials"

### Appendices

#### A Demographic characteristics COVID-19 trials

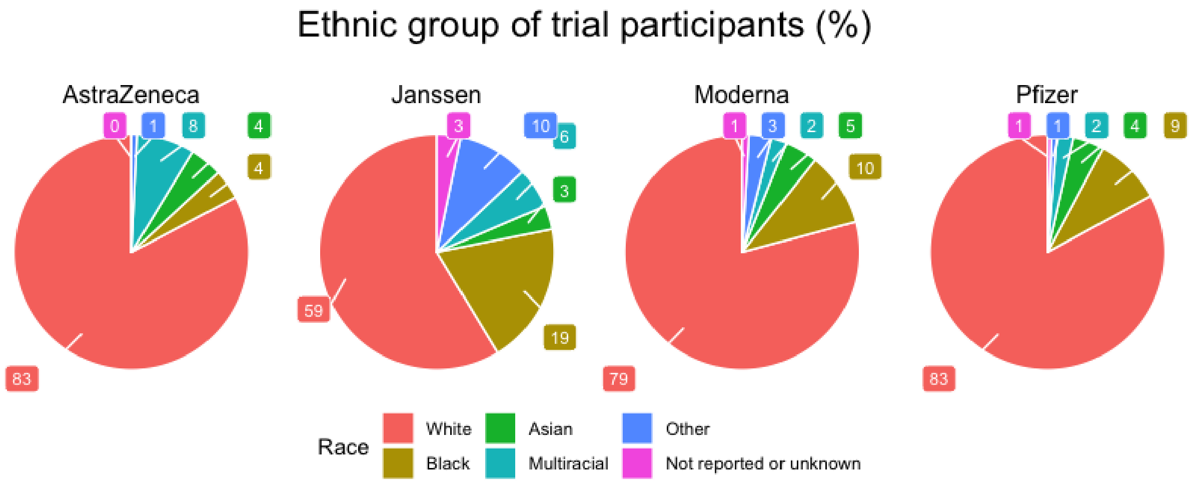


Figure 1: Race distribution of the study participants in the COVID-19 trials conducted by AstraZeneca [32], Janssen [28], Moderna [1] and Pfizer [24].

### B VE risk measure

In this section, we investigate different risk measures for intention-to-treat effects.

#### B.1 Interpretation VE using cumulative incidence

Let $X$ indicate the treatment arm, with $X = 1$ denoting the vaccine arm and $X = 0$ denoting the placebo arm. For a particular patient, let $T^1$ denote the (possibly unobserved) infection time under vaccination and $T^0$ the (possibly unobserved) infection time under placebo. By randomization, $T^x \perp\!\!\!\perp X$ for $x = 0, 1$. Further assuming consistency, stating that the observed infection time corresponds to the potential infection time under the treatment received, i.e. $T^x = T$ for patients with $X = x$, the following inequality then holds for the vaccine efficacy using cumulative incidence as risk measure ($VE_{CI}(t)$):

$$\begin{aligned}
VE_{CI}(t) &= 1 - \frac{P(T \leq t|X = 1)}{P(T \leq t|X = 0)} \\
&= 1 - \frac{P(T^1 \leq t|X = 1)}{P(T^0 \leq t|X = 0)} \\
&= 1 - \frac{P(T^1 \leq t)}{P(T^0 \leq t)} \\
&\leq 1 - \frac{P(T^1 \leq t \cap T^0 \leq t)}{P(T^0 \leq t)} \\
&= P(T^1 > t|T^0 \leq t). \qquad (2)
\end{aligned}$$

Therefore, the vaccine efficacy defined using cumulative incidence can be interpreted as a lower bound for the probability that vaccination prevents infection before a certain time $t$, given that one would be infected by time $t$ if not vaccinated. Consequently, by time $t$, at least 70% of the cases can be avoided by the direct impact of vaccination using a vaccine with 70% efficacy defined using cumulative incidence. In addition, if it is assumed that the infection times under vaccination would always be at least as large as the infection times under placebo, i.e. by making the monotonicity assumption $T^1 \geq T^0$, the inequality in (2) can be replaced by an equality.

#### B.2 Comparison hazard rate and cumulative incidence

We denote the survival curve at time $t$ under treatment $X = x \in \{0, 1\}$ as $S(t|X = x)$, with $X = 0$ denoting the placebo arm and $X = 1$ denoting the vaccine arm. Similarly, the cumulative hazard is noted as $\Lambda(t|X = x)$ and the hazard rate as $\lambda(t|X = x)$. Derivatives with respect to the infection time are denoted as $\Lambda^{'}(t|X = x)$ and $\lambda'(t|X = x)$. The survival function $S(t|X = x)$ can then be approximated using a Taylor expansion at time

$t = 0$:

$$
\begin{aligned}
&S(t|X = x) \\
&= e^{-\Lambda(t|X=x)} \\
&\approx e^{-\Lambda(0|X=x)} - e^{-\Lambda(0|X=x)}\Lambda'(0|X = x)t \\
&+ \left(e^{-\Lambda(0|X=x)}\Lambda'(0|X = x)^2 - e^{-\Lambda(0|X=x)}\Lambda''(0|X = x)\right)\frac{t^2}{2} + \dots \\
&= 1 - \Lambda'(0|X = x)t + \left(\Lambda'(0|X = x)^2 - \Lambda''(0|X = x)\right)\frac{t^2}{2} + \dots \\
&= 1 - \lambda(0|X = x)t + \left(\lambda(0|X = x)^2 - \lambda'(0|X = x)\right)\frac{t^2}{2} + \dots . \qquad (3)
\end{aligned}
$$

The vaccine efficacy at time $t$ defined using the cumulative incidence can be denoted as

$$VE_{CI}(t) := 1 - \frac{P(T \le t|X = 1)}{P(T \le t|X = 0)} = 1 - \frac{1 - S(t|X = 1)}{1 - S(t|X = 0)}.$$

The vaccine efficacy using the hazard ratio is defined

$$VE_{HR}(t) := 1 - \frac{\lambda(t|X = 1)}{\lambda(t|X = 0)} = 1 - e^{\beta},$$

where the second equation assumes proportional hazards, with $\beta$ the log hazard ratio [15].

From (3), it then follows that both definitions are approximately equal up to first order:

$$
\begin{aligned}
VE_{CI}(t) \quad &= 1 - \frac{1 - S(t|X = 1)}{1 - S(t|X = 0)} \\
&\approx 1 - \frac{\lambda(0|X = 1)}{\lambda(0|X = 0)} \\
&= 1 - e^{\beta} \\
&= VE_{HR}(t).
\end{aligned}
$$

Next, we express the VE while approximating the survival curve up to second order:

$$
\begin{aligned}
VE_{CI}(t) \quad &= 1 - \frac{1 - S(t|X = 1)}{1 - S(t|X = 0)} \\
&\approx 1 - \frac{\lambda(0|X = 1)t + \left(\lambda(0|X = 1)^2 - \lambda'(0|X = 1)\right)\frac{t^2}{2}}{\lambda(0|X = 0)t + \left(\lambda(0|X = 0)^2 - \lambda'(0|X = 0)\right)\frac{t^2}{2}} \\
&= 1 - \frac{\lambda(0|X = 0)e^{\beta} + \left(\lambda(0|X = 0)^2 e^{2\beta} - \lambda'(0|X = 0)e^{\beta}\right)\frac{t}{2}}{\lambda(0|X = 0) + \left(\lambda(0|X = 0)^2 - \lambda'(0|X = 0)\right)\frac{t}{2}}.
\end{aligned}
$$

Assuming constant infection hazard under placebo, i.e. $\lambda(t|X = 0) \equiv \lambda_0$, this can be further rewritten as

$$VE_{CI}(t) \quad \approx 1 - e^{\beta}\left(\frac{1+\lambda_0 e^{\beta}\frac{t}{2}}{1+\lambda_0\frac{t}{2}}\right). \tag{4}$$

Therefore, $VE_{CI}(t)$ approximately equals $VE_{HR}(t)$ if $\lambda_0 t \approx 0$, meaning that the cumulative number of cases in the placebo arm is limited. In figure 2, this is illustrated by plotting the differences between $VE_{HR}$ and $VE_{CI}(t)$ over time, for different $VE_{HR}$ values, using expression (4). It can be observed that even for a rather low vaccine efficacy ($VE_{HR} = 0.6$) and cumulative incidence in the placebo arm of 15%, the absolute difference between these vaccine efficacy estimates is only 1.6%. The differences are smaller for higher vaccine efficacy estimates.

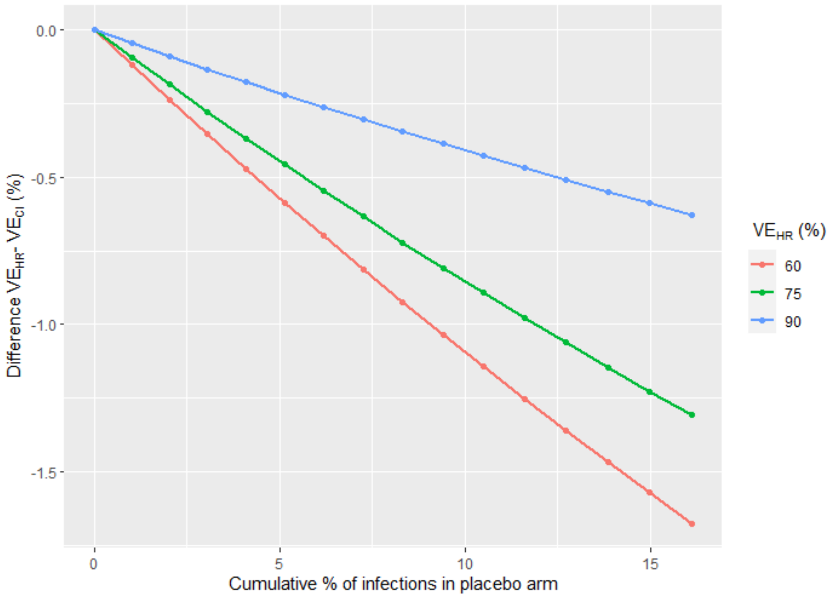


Figure 2: Differences between the VE estimates defined using hazard ratio (HR) and cumulative incidence (CI) in function of the cumulative % of infections in the placebo arm, for a constant infection hazard rate $\lambda_0 = 0.001$.

### C Ramp-up period

#### C.1 ITT versus Per-protocol

In this section, we will express the ITT effect in function of the PP effect where cases observed during the ramp-up period are removed from the analysis, if the vaccine has no effect during this period, i.e. $P(T < \alpha|X = 1) = P(T < \alpha|X = 0)$. We will do this using the cumulative incidence as risk measure.

The ratio of cumulative incidences in the vaccine arm compared to the placebo arm at time $t \geq \alpha$ can be expressed as

$$\begin{aligned}\frac{P(T \leq t|X=1)}{P(T \leq t|X=0)} &= \frac{P(T < \alpha|X=1) + P(T \leq t|X=1, T \geq \alpha)P(T \geq \alpha|X=1)}{P(T < \alpha|X=0) + P(T \leq t|X=0, T \geq \alpha)P(T \geq \alpha|X=0)} \\ &= \frac{P(T < \alpha|X=0) + P(T \leq t|X=1, T \geq \alpha)P(T \geq \alpha|X=0)}{P(T < \alpha|X=0) + P(T \leq t|X=0, T \geq \alpha)P(T \geq \alpha|X=0)}.\end{aligned}$$

Consequently, the ITT effect $VE_{ITT,CI}(t)$ can be expressed in function of the PP effect $VE_{PP,CI}(t)$:

$$\begin{aligned}&VE_{ITT,CI}(t) \\ &= 1 - \frac{P(T \leq t|X=1)}{P(T \leq t|X=0)} \\ &= \left(1 - \frac{P(T \leq t|X=1, T \geq \alpha)}{P(T \leq t|X=0, T \geq \alpha)}\right) \\ &\times \frac{P(T \leq t|X=0, T \geq \alpha)P(T \geq \alpha|X=0)}{P(T < \alpha|X=0) + P(T \leq t|X=0, T \geq \alpha)P(T \geq \alpha|X=0)} \\ &= VE_{PP,CI}(t)\frac{P(T \leq t|X=0, T \geq \alpha)P(T \geq \alpha|X=0)}{P(T < \alpha|X=0) + P(T \leq t|X=0, T \geq \alpha)P(T \geq \alpha|X=0)}. \qquad (5)\end{aligned}$$

Therefore, the size of the difference between the ITT and the PP effect depends on the probability to be infected during the ramp-up time and the probability of becoming infected under placebo before the time point of interest, conditional on not being infected in the ramp-up time.

To investigate how large these differences usually are, expression (5) is calculated for different settings, assuming a constant infection hazard rate. In particular, let $\lambda_0$ denote the infection hazard rate under placebo. It follows that $P(T < \alpha|X = 0) = 1 - e^{-\lambda_0\alpha}$ and $P(T \leq t|X = 0, T \geq \alpha) = 1 - e^{-\lambda_0(t-\alpha)}$. The differences between the two effect measures is then calculated for different infection hazard rates ($\lambda_0 = 0.0001$ and $0.001$) and per-protocol effects ($VE_{PP,CI}(t) = 0.6, 0.75, 0.9$). These differences are calculated at visits $t = 0, 10, 20, \ldots, 150$. In addition, the effect of the length of the ramp-up period is investigated ($\alpha = 7, 14, 28$ days).

In figure 3, it can be seen that the difference between the two vaccine efficacy estimates can be quite large, but decreases as the study progresses. In addition, the difference is slightly larger for larger PP values. As expected, the PP and ITT are closer to each other if the ramp-up period is shorter.

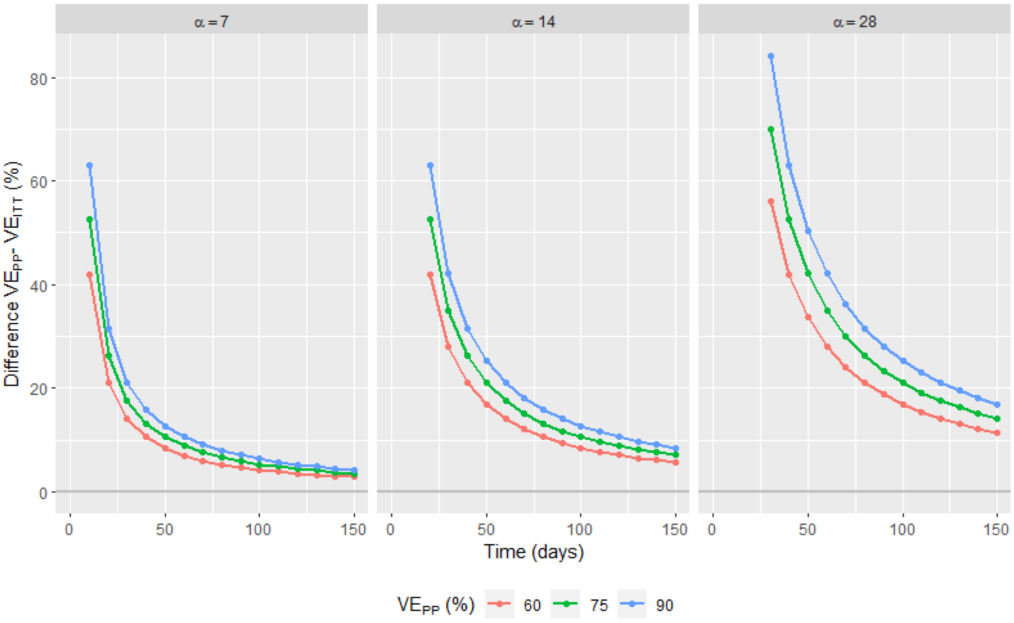


(a) $\lambda_0 = 0.0001$
(Cumulative % of infections in the placebo arm at day 150: 1.5%)

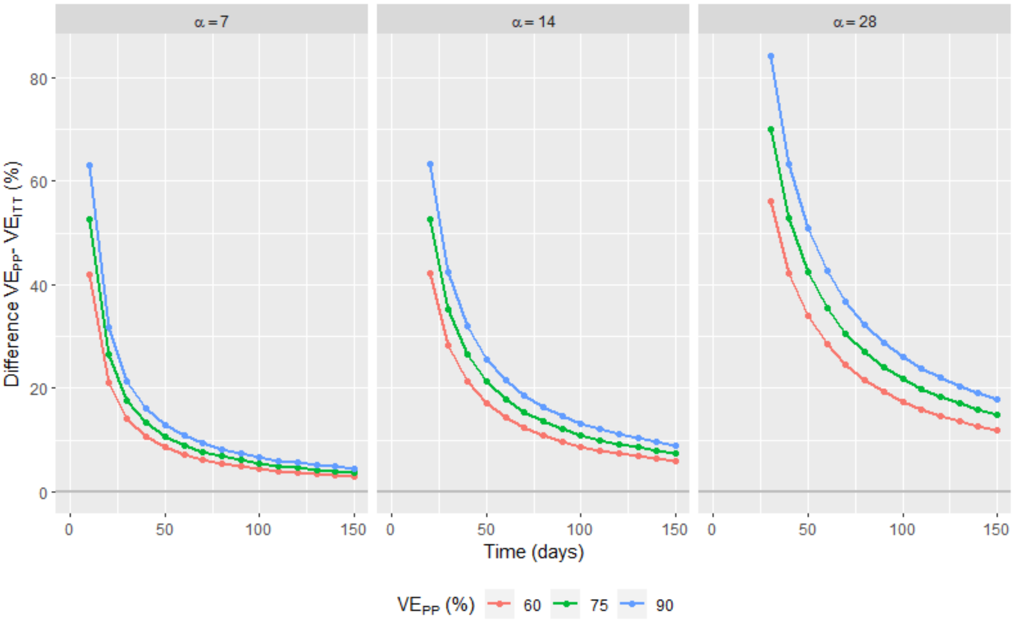


(b) $\lambda_0 = 0.001$
(Cumulative % of infections in the placebo arm at day 150: 14%)

Figure 3: Differences $VE_{PP,CI}(t) - VE_{ITT,CI}(t)$ in function of the time, for a constant infection hazard rate $\lambda_0$. Every point represents the difference in VE that would be obtained if the trial was stopped at the corresponding visit and all information up till that visit was used.

#### C.2 Vaccine efficacy if infections during ramp-up can be prevented

As before, we let $T$ denote the observed infection time for a particular patient and $T^x$ the (possibly unobserved) infection times under vaccination ($x = 1$) or placebo ($x = 0$). Now we also introduce the notation $T^{x0}$, representing the infection times under vaccination ($x = 1$) or placebo ($x = 0$) if infections during the ramp-up time could have been avoided.

When using the cumulative incidence as risk measure, the hypothetical vaccine efficacy estimand (section 3.2.1), i.e. the vaccine efficacy that would have been observed if cases during the ramp-up time could have been avoided, can be expressed as:

$$VE_{\text{hypothetical1},CI}(t) := 1 - \frac{P(T^{10} \leq t)}{P(T^{00} \leq t)}.$$

##### C.2.1 Estimands framework

This estimand can be defined according to the estimands framework described in the addendum of the ICH E9(R1) guideline [19] using the following attributes:

- **Treatment**: experimental vaccine or placebo, as defined by the study protocol.
- **Population**: the entire study population, as defined by the inclusion-exclusion criteria of the study.
- **Variable**: time to infection starting from the day of (first) vaccination.
- **Intercurrent events**: infection during the ramp-up time: the hypothetical scenario is envisaged where infections during the ramp-up time could have been avoided.
- **Population-level summary**: vaccine efficacy defined using cumulative incidence, hazard or incidence rate.

##### C.2.2 Identification

The per-protocol estimator is an unbiased estimator for this estimand under certain (strong) assumptions. In particular, assume that patients who are infected during the ramp-up time ($T^x < \alpha$) are comparable to patients who were not infected during this period in terms of their counterfactual infection times if infections during the ramp-up time could have been avoided, i.e.

$$P(T^{x0} \leq t | T^x < \alpha) = P(T^{x0} \leq t | T^x \geq \alpha) \quad \forall t, \tag{6}$$

and consequently $P(T^{x0} \leq t) = P(T^{x0} \leq t | T^x \geq \alpha)$. From this assumption it follows that

$$\begin{aligned} VE_{\text{Hypothetical1},CI}(t) &= 1 - \frac{P(T^{10} \leq t)}{P(T^{00} \leq t)} \\ &= 1 - \frac{P(T^{10} \leq t | T^1 \geq \alpha)}{P(T^{00} \leq t | T^0 \geq \alpha)}. \end{aligned} \tag{7}$$

Next, assume that $T^x = T^{x0}$ for patients who were not infected during the ramp-up time. From this assumption, expression (7) can be rewritten as

$$\begin{aligned} VE_{\text{Hypothetical1},CI}(t) &= 1 - \frac{P(T^1 \leq t | T^1 \geq \alpha)}{P(T^0 \leq t | T^0 \geq \alpha)} \\ &= 1 - \frac{P(T \leq t | X = 1, T \geq \alpha)}{P(T \leq t | X = 0, T \geq \alpha)} \\ &= VE_{PP,CI}(t), \end{aligned} \tag{8}$$

where the second equality holds by randomization ($T^x \perp\!\!\!\perp X$ for $x = 0, 1$), and assuming consistency, stating that the observed infection time corresponds to the potential infection time under the treatment received, i.e. $T^x = T$ for patients with $X = x$.

Assumption (6) is a strong assumption since patients who are infected during the ramp-up time may have a higher risk of becoming infected quickly. The assumption that infection times of patients who were not infected during the ramp-up time are not affected by the intervention, i.e. $T^x = T^{x0}$, is also strong. In addition, the effect of the infections during the ramp-up period on the infection times of other patients is ignored.

#### C.3 Vaccine efficacy if ramp-up period can be eliminated

In this section, we discuss an estimand and estimator to estimate the hypothetical vaccine efficacy as proposed in section 3.2.2. For this, we use an approach inspired by structural accelerated failure time (AFT) models [27, 16]. First, we show how this estimand can be defined according to the estimands framework described in the addendum of the ICH E9(R1) guideline [19]. Next, we review structural AFT models to model infection times. Afterwards, we propose our method.

##### C.3.1 Estimands framework

This estimand can be defined according to the estimands framework described in the addendum of the ICH E9(R1) guideline [19] using the following attributes:

- **Treatment**: experimental vaccine or placebo, as defined by the study protocol.
- **Population**: the entire study population, as defined by the inclusion-exclusion criteria of the study.

- **Variable**: time to infection starting from the day of (first) vaccination.
- **Intercurrent events**: infection during the ramp-up time: the hypothetical scenario is envisaged where the ramp-up period could have been eliminated.
- **Population-level summary**: vaccine efficacy defined using cumulative incidence, hazard or incidence rate.

##### C.3.2 Structural AFT models

Structural accelerated failure time models [27, 16] can be used to model the infection times under vaccine $T^1$ in function of the infection times under placebo $T^0$. In particular, it is assumed that

$$T^1 = \int_0^{T^0} \exp(\Psi(t))dt, \tag{9}$$

for a function $\Psi(t)$ that represents the causal effect of the vaccine on the time to infection. If the parameter $\alpha$ denotes the length of the ramp-up period, then one could for example use the function

$$\Psi(t) = \begin{cases} \rho\psi & \text{if } t < \alpha/\exp(\rho\psi) \\ \psi & \text{if } t \geq \alpha/\exp(\rho\psi), \end{cases} \tag{10}$$

for scalar values $\rho$ and $\psi$, leading to

$$T^1 = \begin{cases} \exp(\rho\psi)T^0 & \text{if } T^0 < \alpha/\exp(\rho\psi) \\ \exp(\psi)T^0 + \alpha(1 - \exp(\psi(1-\rho))) & \text{if } T^0 \geq \alpha/\exp(\rho\psi). \end{cases} \tag{11}$$

In this model, it is assumed that vaccination extends or shortens the infection times by a certain factor, depending on the parameter values $\psi$, $\rho$ and $\alpha$. In particular, $\psi$ represents the vaccine effect, with higher values indicating higher efficacy. The value $\psi = 0$ encodes the strong null hypothesis of no vaccine effect on the infection times, i.e. $T^1 = T^0$ for all patients. Parameter $\rho$ indicates how much weaker the vaccine effect is during the ramp-up time than after the ramp-up time. The choice $\rho = 0$ represents the assumption of no vaccine effect during the ramp-up period, while $\rho = 1$ indicates full vaccine effect from baseline.

The boundaries in function (10) are chosen to ensure that the full vaccine effect starts at the end of the ramp-up period, i.e. on day $\alpha$. In particular, let $S^1(t)$ and $S^0(t)$ denote the infection survival functions under vaccine and under placebo, respectively. These functions are linked to each other in the following way:

$$
\begin{aligned}
&S^1(t) \\
&= P(T^1 \geq t) \\
&= P(\exp(\rho\psi)T^0 \geq t | T^0 < \alpha / \exp(\rho\psi)) P(T^0 < \alpha / \exp(\rho\psi)) \\
&+ P(\exp(\psi)T^0 + \alpha(1 - \exp(\psi(1-\rho))) \geq t | T^0 \geq \alpha / \exp(\rho\psi)) P(T^0 \geq \alpha / \exp(\rho\psi)) \\
&= P\left(\frac{t}{\exp(\rho\psi)} \leq T^0 < \frac{\alpha}{\exp(\rho\psi)}\right) + P\left(T^0 \geq \max\left\{\frac{t - \alpha(1-\exp(\psi(1-\rho)))}{\exp(\psi)}, \frac{\alpha}{\exp(\rho\psi)}\right\}\right) \\
&= \left\{P\left(T^0 < \frac{\alpha}{\exp(\rho\psi)}\right) - P\left(T^0 < \frac{t}{\exp(\rho\psi)}\right)\right\} I(t < \alpha) \\
&+ P\left(T^0 \geq \frac{t - \alpha(1-\exp(\psi(1-\rho)))}{\exp(\psi)}\right) I(t \geq \alpha) + P\left(T^0 \geq \frac{\alpha}{\exp(\rho\psi)}\right) I(t < \alpha) \\
&= \left\{1 - P\left(T^0 < \frac{t}{\exp(\rho\psi)}\right)\right\} I(t < \alpha) + P\left(T^0 \geq \frac{t - \alpha(1-\exp(\psi(1-\rho)))}{\exp(\psi)}\right) I(t \geq \alpha) \\
&= \begin{cases} P\left(T^0 \geq \frac{t}{\exp(\rho\psi)}\right) & \text{if } t < \alpha \\ P\left(T^0 \geq \frac{t-\alpha(1-\exp(\psi(1-\rho)))}{\exp(\psi)}\right) & \text{if } t \geq \alpha \end{cases} \\
&= \begin{cases} S^0\left(\frac{t}{\exp(\rho\psi)}\right) & \text{if } t < \alpha \\ S^0\left(\frac{t-\alpha(1-\exp(\psi(1-\rho)))}{\exp(\psi)}\right) & \text{if } t \geq \alpha. \end{cases}
\end{aligned}
$$

With this parameterization we thus establish that the vaccine has a different effect on the survival function before and after $t = \alpha$. This model is visualized in figure 4.

Model (9) is deterministic, in the sense that it assumes that, for each subject, the counterfactual infection time under vaccination can be calculated without error as a function of $T^0$ and $\Psi(t)$. In addition, this model assumes rank preservation for the patients' infection times across treatment regimes [16], which is seldom plausible [31]. Moreover, available methods for fitting rank preserving structural failure time models cannot easily accommodate censoring of survival times, since infection times under vaccination cannot be calculated using model (10) when the corresponding infection time under placebo is censored [20]. In the next section, these disadvantages will be tackled, by transforming population survival functions instead of individual infection times.

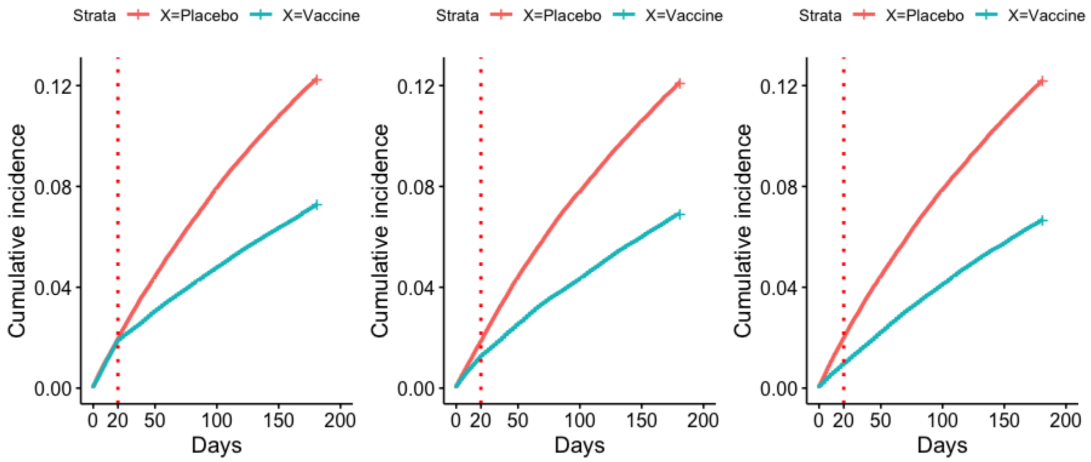


(a) $\rho = 0$ (b) $\rho = 0.6$ (c) $\rho = 1$

Figure 4: Cumulative incidence plots for infection times, illustrating structural AFT model (11) or SDM (12), with $\alpha = 20$ days and $\psi = 0.8$, for different $\rho$ values.

##### C.3.3 Structural Distribution Models

Instead of making a mapping between the potential infection times themselves using structural AFT models, we will map percentiles of the distribution of infection times under placebo into percentiles of the distribution of infection times under vaccine using a Structural Distribution Model (SDM) [26, 31].

First, let $S(t|X = 1)$ and $S(t|X = 0)$ denote the survival probabilities at time $t$ in the vaccine and placebo arm. Next, the survival function in the vaccine arm is mapped into the survival function in the placebo arm. In particular, if $S_{\text{SDM}}(t|X = 1)$ represents the mapped survival function in the vaccine arm, it is postulated that

$$S_{\text{SDM}}(t|X = 1) = S(\gamma(t; \Phi)|X = 0),$$

for all $t$, with $\gamma(t; \Phi)$ a known function satisfying $\gamma(0; \Phi) = 0$. Here, we use the mapping between the infection times in model (11) to determine $\gamma(t;\Phi)$:

$$\begin{aligned}
&S_{\mathrm{SDM}}(t|X=1;\rho,\psi,\alpha)\\
&=P(T\geq t|X=1;\rho,\psi,\alpha)\\
&=P(T^1\geq t;\rho,\psi,\alpha)\\
&=P(\exp(\rho\psi)T^0\geq t|T^0<\alpha/\exp(\rho\psi))P(T^0<\alpha/\exp(\rho\psi))\\
&+P(\exp(\psi)T^0+\alpha(1-\exp(\psi(1-\rho)))\geq t|T^0\geq\alpha/\exp(\rho\psi))P(T^0\geq\alpha/\exp(\rho\psi))\\
&=P\left(\frac{t}{\exp(\rho\psi)}\leq T^0<\frac{\alpha}{\exp(\rho\psi)}\right)+P\left(T^0\geq\max\left\{\frac{t-\alpha(1-\exp(\psi(1-\rho)))}{\exp(\psi)},\frac{\alpha}{\exp(\rho\psi)}\right\}\right)\\
&=\left\{P\left(T^0<\frac{\alpha}{\exp(\rho\psi)}\right)-P\left(T^0<\frac{t}{\exp(\rho\psi)}\right)\right\}I(t<\alpha)\\
&+P\left(T^0\geq\frac{t-\alpha(1-\exp(\psi(1-\rho)))}{\exp(\psi)}\right)I(t\geq\alpha)+P\left(T^0\geq\frac{\alpha}{\exp(\rho\psi)}\right)I(t<\alpha)\\
&=\left\{1-P\left(T^0<\frac{t}{\exp(\rho\psi)}\right)\right\}I(t<\alpha)+P\left(T^0\geq\frac{t-\alpha(1-\exp(\psi(1-\rho)))}{\exp(\psi)}\right)I(t\geq\alpha)\\
&=S\left(\frac{t}{\exp(\rho\psi)}|X=0\right)I(t<\alpha)+S\left(\frac{t-\alpha(1-\exp(\psi(1-\rho)))}{\exp(\psi)}|X=0\right)I(t\geq\alpha),
\end{aligned}\tag{12}$$

with $I(.)$ the indicator function and assuming consistency, i.e. $T^x=T$ for patients with $X=x$. From this, it follows that

$$\gamma(t;\rho,\psi,\alpha)=\frac{t}{\exp(\rho\psi)}I(t<\alpha)+\frac{t-\alpha(1-\exp(\psi(1-\rho)))}{\exp(\psi)}I(t\geq\alpha).$$

In this parametrisation, $\rho=0$ represents the assumption of no vaccine effect during the ramp-up period, while $\rho=1$ indicates full vaccine effect from baseline. A value $\rho\in]0,1[$ parameterizes a limited effect during the ramp-up time, as illustrated in figure 5. If $\psi$ would be set to 0, this survival curve under vaccination would simplify to the placebo survival curve, i.e. $S_{\mathrm{SDM}}(t|X=1;\rho,\psi=0,\alpha)=S(t|X=0)$, encoding the null hypothesis of no vaccine effect on the infection times. A higher value of $\psi$ indicates a higher vaccine efficacy, as illustrated in figure 6. Distribution model (12) assumes that vaccination shifts each percentile of the distribution of $T$ by a value $\gamma^*(\rho,\psi,\alpha)$ constant for all $t$, i.e. $\gamma(t;\rho,\psi,\alpha)=t-\gamma^*(\rho,\psi,\alpha)$ [31].

If the vaccine has no effect at all during the ramp-up time, i.e. $\rho=0$, model (12) simplifies to

$$S_{\mathrm{SDM}}(t|X=1;\rho=0,\psi,\alpha)=\begin{cases}S(t|X=0) & \text{if } t<\alpha\\ S\left(\frac{t-\alpha}{\exp(\psi)}+\alpha|X=0\right) & \text{if } t\geq\alpha,\end{cases}$$

from which it can be seen that the survival curves in the two arms coincide during the ramp-up time. After the ramp-up time, the vaccine is fully effective and the quantiles of the infection distribution are multiplied by a factor $\exp(\psi)$. However, the period in which the vaccine was not yet effective needs to be taken into account, hence the extra term $\alpha$ in the survival function. If the vaccine has already an effect during the ramp-up time, i.e. $\rho \in ]0, 1]$, model (12) can be rewritten as

$$S_{\text{SDM}}(t|X=1;\rho,\psi,\alpha) = \begin{cases} S\left(\frac{t}{\exp(\rho\psi)}|X=0\right) & \text{if } t < \alpha \\ S\left(\frac{t-\alpha}{\exp(\psi)} + \frac{\alpha}{\exp(\rho\psi)}|X=0\right) & \text{if } t \geq \alpha. \end{cases}$$

During the ramp-up time, the quantiles of the infection distribution are not multiplied by $\exp(\psi)$, but by a smaller factor $\exp(\rho\psi)$, encoding the fact that the vaccine is not fully effective yet. After the ramp-up time, the vaccine is fully effective and the quantiles are multiplied by $\exp(\psi)$, but the ramp-up period needs to be taken into account, hence the extra term $\alpha/\exp(\rho\psi)$ in the survival function.

The survival probabilities $S(t|X=1)$ and $S(t|X=0)$ can be estimated by Kaplan-Meier estimators [21]. The censored infection times in SDM (12) are then automatically handled by the Kaplan-Meier estimator. Estimation of the parameters $\rho, \psi$ and $\alpha$ is discussed in the next section.

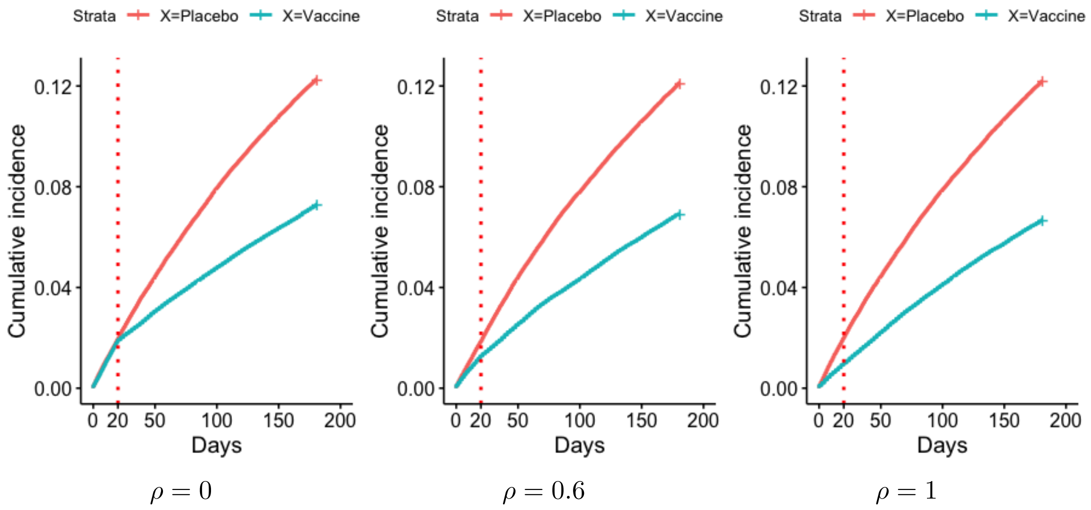


Figure 5: Illustration of parameter $\rho$ in model (12) for fixed $\psi = 0.8$ and $\alpha = 20$.

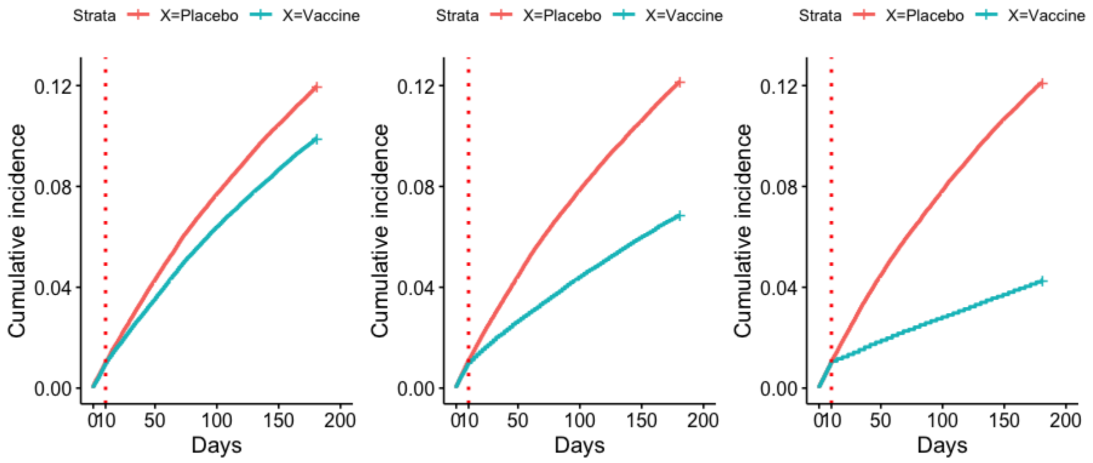


$\psi = 0.3$ $\psi = 0.8$ $\psi = 1.5$

Figure 6: Illustration of parameter $\psi$ in model (12) for fixed $\alpha = 10$ and $\rho = 0$.

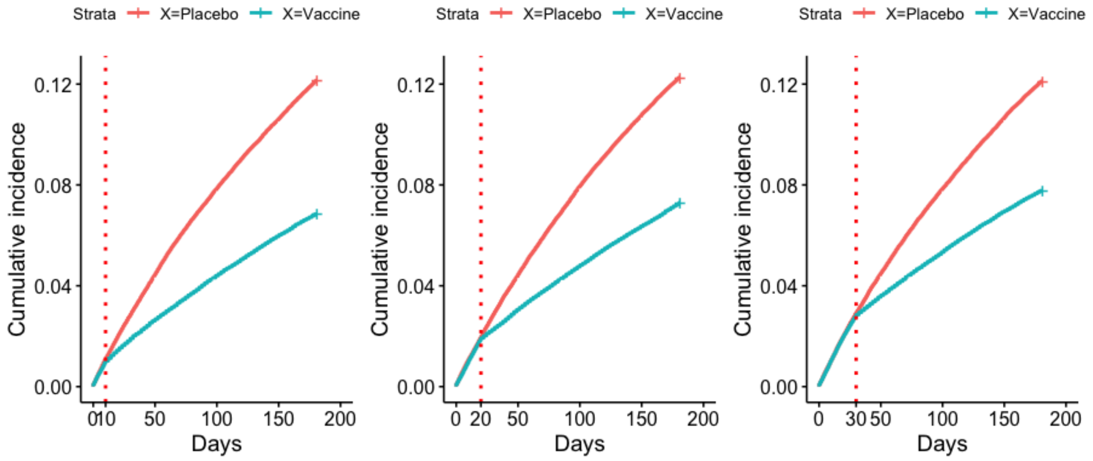


$\alpha = 10$ $\alpha = 20$ $\alpha = 30$

Figure 7: Illustration of parameter $\alpha$ in model (12) for fixed $\rho = 0$ and $\psi = 0.8$.

##### C.3.4 Estimating parameters

The modeled survival curve $S_{\text{SDM}}(t|X = 1; \rho, \psi, \alpha)$ (12) can be compared to the observed Kaplan-Meier survival curve in the vaccine arm $\hat{S}(t|X = 1)$ to obtain estimates for $\rho, \psi$ and $\alpha$. However, it may be too ambitious to estimate all of these parameters. Therefore, we propose three approaches whereby 1, 2 or 3 of these parameters are estimated, while the others are specified by the user based on knowledge about the vaccine. R-code for these methods is provided in Appendix D.2.4.

**Method 1: Only estimating $\psi$**

Let $t = L$ denote the end of the trial, i.e. the day at which the primary analysis is performed. The following steps can be used to estimate $\psi$ in model (12), while the length of the ramp-up period, $\alpha$, and the parameter for the effectiveness of the vaccine during the ramp-up period, $\rho$, are pre-specified based on the knowledge about how the vaccine works:

1. Estimate Kaplan-Meier estimators for the survival probabilities in the vaccine and placebo arm. Let $\hat{S}(t|X = 1)$ and $\hat{S}(t|X = 0)$ denote the obtained estimators.

2. Obtain estimates for $\psi$ in model (12) by comparing the mapped survival function

$$S_{\mathrm{SDM}}(t|X = 1; \rho, \hat{\psi}, \alpha)$$
$$:= \hat{S}\left(\frac{t}{\exp(\rho\hat{\psi})}|X = 0\right) I(t < \alpha) + \hat{S}\left(\frac{t - \alpha(1 - \exp(\hat{\psi}(1 - \rho)))}{\exp(\hat{\psi})}|X = 0\right) I(t \geq \alpha),$$

   to the Kaplan-Meier estimator $\hat{S}(t|X = 1)$:

   (a) First, an initial estimate for $\hat{\psi}$ is made by minimizing the squared difference in survival probabilities at the end of the trial:

$$\left(S_{\mathrm{SDM}}(L|X = 1; \rho, \hat{\psi}, \alpha) - \hat{S}(L|X = 1)\right)^2. \tag{13}$$

   This squared difference can be calculated for a range of values for $\hat{\psi}$, e.g. $\{0, 0.1, 0.2, \ldots, 10\}$. Let $\hat{\psi}_0$ denote the value in this interval for which the squared difference (13) is minimized.

   (b) Second, this initial estimate $\hat{\psi}_0$ is updated by minimizing the squared difference in restricted mean survival times over the entire duration of the trial:

$$\left(\int_0^L S_{\mathrm{SDM}}(t|X = 1; \rho, \hat{\psi}, \alpha)dt - \int_0^L \hat{S}(t|X = 1)dt\right)^2. \tag{14}$$

   Here, $\hat{\psi}$ denotes the value for $\psi$ for which this squared difference is minimized. The estimator $\hat{\psi}_0$ can be specified as starting value for the minimization.

The standard errors of the estimator for $\psi$ can be obtained by performing non-parametric bootstrap replications [8].

**Method 2: Estimating $\psi$ and $\rho$**

The following steps can be used to estimate $\psi$ and $\rho$ in model (12), while the length of the ramp-up period, $\alpha$, is pre-specified based on the knowledge about the vaccine:

1. Estimate Kaplan-Meier estimators for the survival probabilities in the vaccine and placebo arm. Let $\hat{S}(t|X=1)$ and $\hat{S}(t|X=0)$ denote the obtained estimators.

2. Obtain estimates for $\psi$ and $\rho$ in model (12) by comparing the mapped survival function

$$S_{\text{SDM}}(t|X=1;\hat{\rho},\hat{\psi},\alpha)$$
$$:=\hat{S}\left(\frac{t}{\exp(\hat{\rho}\hat{\psi})}|X=0\right)I(t<\alpha)+\hat{S}\left(\frac{t-\alpha(1-\exp(\hat{\psi}(1-\hat{\rho})))}{\exp(\hat{\psi})}|X=0\right)I(t\geq\alpha),$$

with $\hat{\rho}$ and $\hat{\psi}$ estimators for $\rho$ and $\psi$, to the Kaplan-Meier estimator $\hat{S}(t|X=1)$:

   (a) First, initial estimates for $\hat{\psi}$ and $\hat{\rho}$ are made by minimizing the sum of squared differences in survival probabilities at $t=\alpha$ and $t=L$:

$$\left(S_{\text{SDM}}(\alpha|X=1;\hat{\rho},\hat{\psi},\alpha)-\hat{S}(\alpha|X=1)\right)^2$$
$$+\left(S_{\text{SDM}}(L|X=1;\hat{\rho},\hat{\psi},\alpha)-\hat{S}(L|X=1)\right)^2. \tag{15}$$

   This sum of the squared differences can be calculated for a range of values for $\hat{\psi}$, e.g. $\{0,0.5,1,\ldots,10\}$, and for $\hat{\rho}$, e.g. $\{0,0.1,0.2,\ldots,1\}$. Let $\hat{\psi}_0$ and $\hat{\rho}_0$ denote the values for $\hat{\psi}$ and $\hat{\rho}$ for which expression (15) is minimized.

   (b) Second, these initial estimates $\hat{\psi}_0$ and $\hat{\rho}_0$ are updated by minimizing sum of squared differences in restricted mean survival times

$$\left(\int_0^{L/2}S_{\text{SDM}}(t|X=1;\hat{\rho},\hat{\psi},\alpha)dt-\int_0^{L/2}\hat{S}(t|X=1)dt\right)^2$$
$$+\left(\int_0^{L}S_{\text{SDM}}(t|X=1;\hat{\rho},\hat{\psi},\alpha)dt-\int_0^{L}\hat{S}(t|X=1)dt\right)^2. \tag{16}$$

   Here, $\hat{\rho}$ and $\hat{\psi}$ denote the values for $\rho$ and $\psi$ for which this squared difference is minimized. The estimators $\hat{\psi}_0$ and $\hat{\rho}_0$ can be specified as starting values for the minimization.

The standard errors of the estimator for $\psi$ and $\rho$ can be obtained by performing non-parametric bootstrap replications [8].

**Method 3: Estimating $\psi$, $\rho$ and $\alpha$**

The following steps can be used to estimate $\psi$, $\rho$ and $\alpha$ in model (12):

1. Estimate Kaplan-Meier estimators for the survival probabilities in the vaccine and placebo arm. Let $\hat{S}(t|X=1)$ and $\hat{S}(t|X=0)$ denote the obtained estimators.

2. Obtain estimates for $\psi$ and $\rho$ in model (12) by comparing the mapped survival function

$$S_{\mathrm{SDM}}(t|X=1;\hat{\rho},\hat{\psi},\hat{\alpha})$$
$$:= \hat{S}\left(\frac{t}{\exp(\hat{\rho}\hat{\psi})}|X=0\right) I(t<\hat{\alpha}) + \hat{S}\left(\frac{t-\hat{\alpha}(1-\exp(\hat{\psi}(1-\hat{\rho})))}{\exp(\hat{\psi})}|X=0\right) I(t\geq\hat{\alpha}),$$

with $\hat{\rho}$, $\hat{\psi}$ and $\hat{\alpha}$ estimators for $\rho$, $\psi$ and $\alpha$, to the Kaplan-Meier estimator $\hat{S}(t|X=1)$:

   (a) First, initial estimates for $\hat{\psi}$, $\hat{\rho}$ and $\hat{\alpha}$ are made by minimizing the sum of squared differences in survival probabilities at $t = L/4$, $t = L/2$ and $t = L$:

$$\begin{aligned}&\Big(S_{\mathrm{SDM}}(L/4|X=1;\hat{\rho},\hat{\psi},\hat{\alpha}) - \hat{S}(L/4|X=1)\Big)^2\\ &+\Big(S_{\mathrm{SDM}}(L/2|X=1;\hat{\rho},\hat{\psi},\hat{\alpha}) - \hat{S}(L/2|X=1)\Big)^2\\ &+\Big(S_{\mathrm{SDM}}(L|X=1;\hat{\rho},\hat{\psi},\hat{\alpha}) - \hat{S}(L|X=1)\Big)^2. \qquad (17)\end{aligned}$$

   This sum of the squared differences can be calculated for a range of values for $\hat{\psi}$, e.g. $\{0, 0.5, 1, \ldots, 10\}$, for $\hat{\rho}$, e.g. $\{0, 0.1, 0.2, \ldots, 1\}$ and for $\hat{\alpha}$, e.g. $\{0, 1, 2, \ldots, L/2\}$. Let $\hat{\psi}_0$, $\hat{\rho}_0$ and $\hat{\alpha}_0$ denote the values for $\hat{\psi}$, $\hat{\rho}$ and $\hat{\alpha}$ for which expression (17) is minimized.

   (b) Second, these initial estimates $\hat{\psi}_0$, $\hat{\rho}_0$ and $\hat{\alpha}_0$ are updated by minimizing sum of squared differences in restricted mean survival times

$$\begin{aligned}&\left(\int_0^{L/4} S_{\mathrm{SDM}}(t|X=1;\hat{\rho},\hat{\psi},\hat{\alpha})dt - \int_0^{L/4}\hat{S}(t|X=1)dt\right)^2\\ &+\left(\int_0^{L/2} S_{\mathrm{SDM}}(t|X=1;\hat{\rho},\hat{\psi},\hat{\alpha})dt - \int_0^{L/2}\hat{S}(t|X=1)dt\right)^2\\ &+\left(\int_0^{L} S_{\mathrm{SDM}}(t|X=1;\hat{\rho},\hat{\psi},\hat{\alpha})dt - \int_0^{L}\hat{S}(t|X=1)dt\right)^2. \qquad (18)\end{aligned}$$

   Here, $\hat{\rho}$, $\hat{\psi}$ and $\hat{\alpha}$ denote the values for $\rho$, $\psi$ and $\alpha$ for which this squared difference is minimized. The estimators $\hat{\psi}_0$, $\hat{\rho}_0$ and $\hat{\alpha}_0$ can be specified as starting values for the minimization.

The standard errors of the estimators for $\psi$, $\rho$ and $\alpha$ can be obtained by performing nonparametric bootstrap replications [8].

##### C.3.5 Estimating vaccine efficacy if ramp-up period can be eliminated

After obtaining estimates or specifying values for $\rho, \psi$ and $\alpha$, the hypothetical estimand can be obtained by setting the ramp-up period to 0 days in model (12):

$$\begin{aligned}\widehat{VE}_{\text{Hypothetical2},CI}(t) \quad &= 1 - \frac{1 - S_{\text{SDM}}(t|X=1;\rho,\psi,\alpha=0)}{1-\hat{S}(t|X=0)} \\ &= 1 - \frac{1-\hat{S}(t/\exp(\psi)|X=0)}{1-\hat{S}(t|X=0)}, \end{aligned} \tag{19}$$

with $\hat{S}(t|X = 0)$ the Kaplan-Meier estimator for the survival function in the placebo arm. This VE estimator (19) relies on the assumption that the mapped survival curve $S_{\text{SDM}}(t|X = 1;\rho,\psi,\alpha)$ is correct. In addition, it is assumed that this survival curve can be extrapolated to the setting with no ramp-up time. The standard errors of this vaccine efficacy estimator can be obtained by performing non-parametric bootstrap replications [8].

#### C.4 Data analysis

##### C.4.1 Data

The data of the Janssen and Pfizer COVID-19 trials were recreated, based on the published Kaplan-Meier curves (Figure 1 in [13] and Figure 2 in [12]). In the Janssen trial, approximately 40000 patients were randomized, while the Pfizer trial included approximately 44000 patients. In the Janssen trial, a total of 625 cases were observed (193 in the placebo arm and 432 in the vaccine arm), from which 464 at least 14 days after vaccination (116 in the placebo arm and 348 in the vaccine arm). In the Pfizer trial, 325 cases were observed (50 in the placebo arm and 275 in the vaccine arm), from which 170 with onset at least 7 days after the second dose (8 in the placebo arm and 162 in the vaccine arm).

Both companies reported cumulative incidence curves, together with the number of events and patients at risk per 7 days. Based on these numbers, the data was recreated, whereby patients could be infected or censored at every day of the trial. In particular, the reported infected and censored cases per week were randomly divided over that week, resulting in a dataset where the same number of patients were randomized, infected and censored as in the real trial. The simulated data is shown in figures 8 and 9 in Appendix C.4. The R-code for producing these datasets is provided in Appendix D.1.

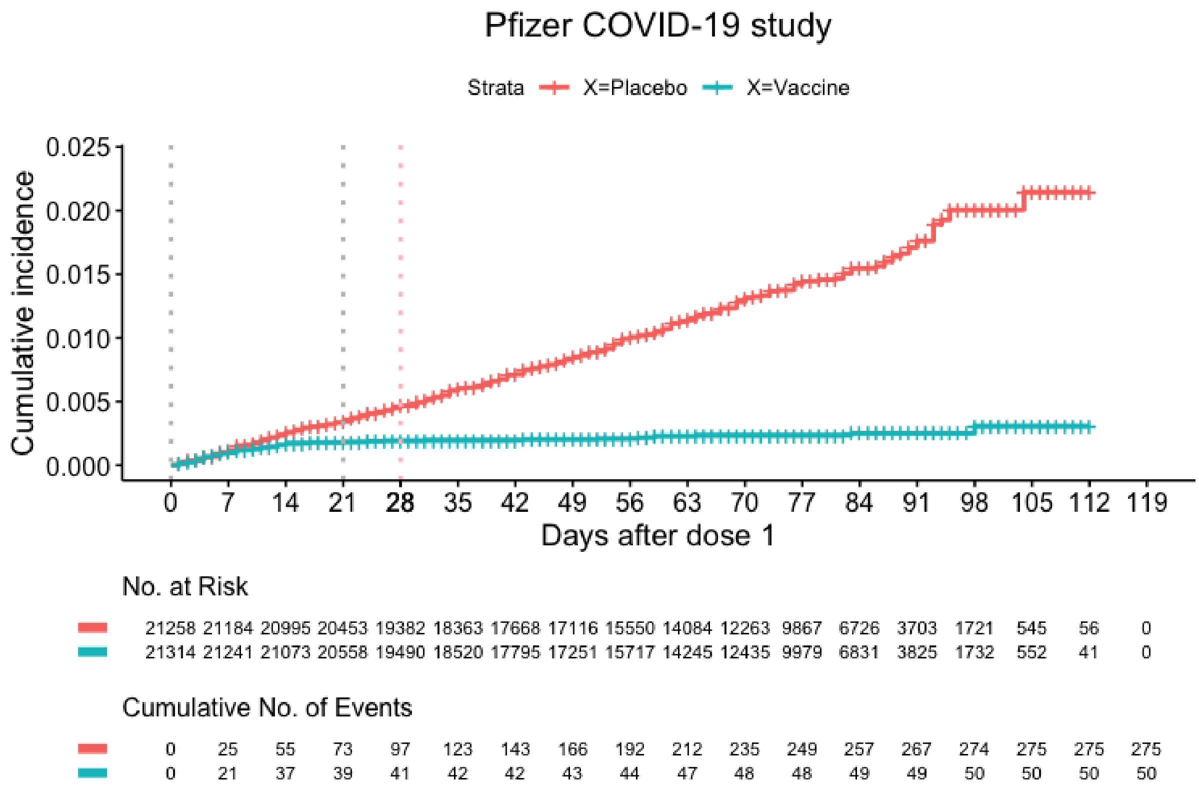


Figure 8: Simulated data similar to the COVID-19 vaccine trial conducted by Pfizer [12]. The grey vertical lines indicate the visits at which a dose of the vaccine is given and the pink line indicates the end of the ramp-up period.
Note that 'No. at Risk' represents the number of patients at risk before the corresponding visit (not including that corresponding visit). Therefore, these values do not exactly coincide with the numbers stated in the Pfizer briefing document.

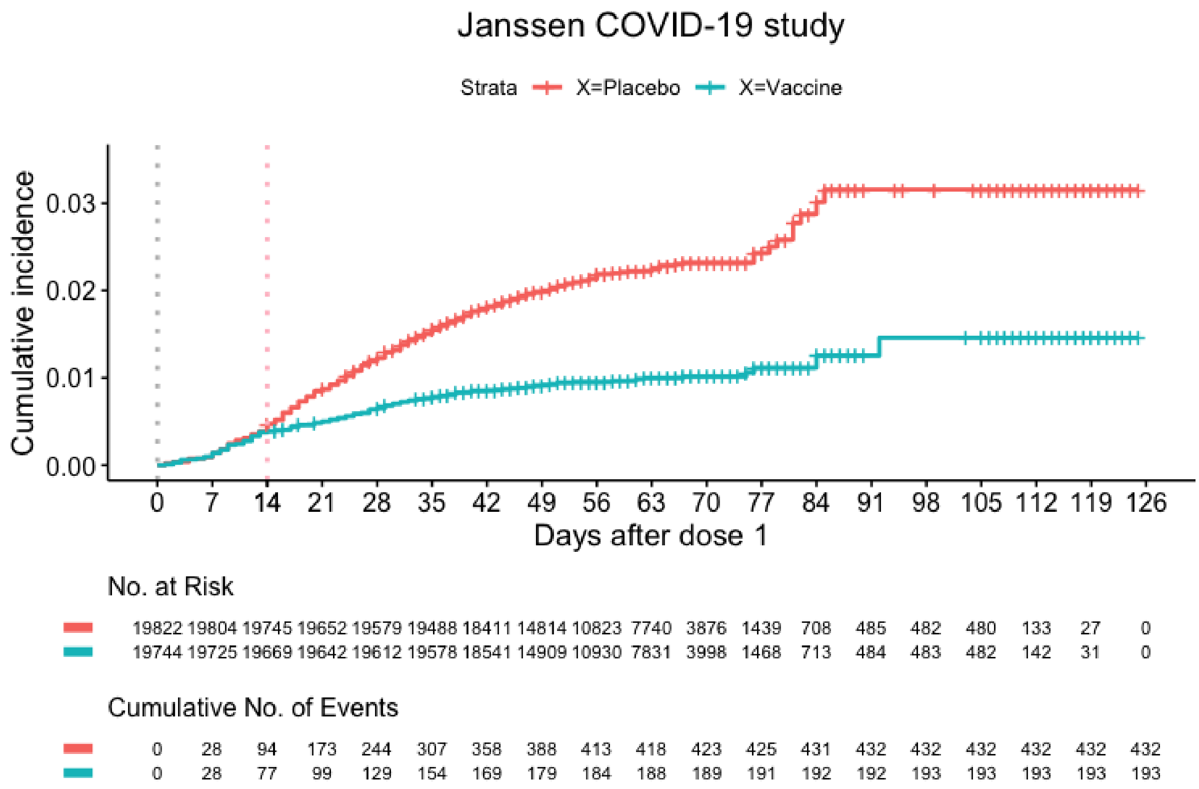


Figure 9: Simulated data similar to the COVID-19 vaccine trial conducted by Janssen [13]. The grey vertical line indicates the visit at which a dose of the vaccine is given and the pink line indicates the end of the ramp-up period.

##### C.4.2 Results ITT versus Per-protocol

| **Ramp-up period** | **Effect** | **Risk measure** | **VE (SE)** |
|---|---|---|---|
| | Intention-to-treat | Hazard rate | 0.82 (0.03) |
| | | Cumulative incidence | 0.86 (0.03) |
| | | Incidence rate | 0.82 (0.03) |
| $\boldsymbol{\alpha = 7}$ **days** | Per-protocol | Hazard rate | 0.88 (0.02) |
| cases before $\alpha$: | (removing cases before $\alpha$) | Cumulative incidence | 0.90 (0.03) |
| 8 % of placebo cases | | Incidence rate | 0.88 (0.02) |
| 40% of vaccine cases | Per-protocol | Hazard rate | 0.88 (0.02) |
| | (censoring cases before $\alpha$) | Cumulative incidence | 0.90 (0.04) |
| | | Incidence rate | 0.88 (0.02) |
| $\boldsymbol{\alpha = 14}$ **days** | Per-protocol | Hazard rate | 0.93 (0.02) |
| cases before $\alpha$: | (removing cases before $\alpha$) | Cumulative incidence | 0.92 (0.04) |
| 18 % of placebo cases | | Incidence rate | 0.93 (0.02) |
| 68 % of vaccine cases | Per-protocol | Hazard rate | 0.93 (0.02) |
| | (censoring cases before $\alpha$) | Cumulative incidence | 0.92 (0.03) |
| | | Incidence rate | 0.93 (0.02) |
| $\boldsymbol{\alpha = 28}$ **days** | Per-protocol | Hazard rate | 0.95 (0.02) |
| cases before $\alpha$: | (removing cases before $\alpha$) | Cumulative incidence | 0.93 (0.04) |
| 34 % of placebo cases | | Incidence rate | 0.95 (0.02) |
| 82 % of vaccine cases | Per-protocol | Hazard rate | 0.95 (0.02) |
| | (censoring cases before $\alpha$) | Cumulative incidence | 0.93 (0.04) |
| | | Incidence rate | 0.95 (0.02) |
| $\boldsymbol{\alpha = 35}$ **days** | Per-protocol | Hazard rate | 0.95 (0.02) |
| cases before $\alpha$: | (removing cases before $\alpha$) | Cumulative incidence | 0.93 (0.04) |
| 44 % of placebo cases | | Incidence rate | 0.95 (0.02) |
| 84 % of vaccine cases | Per-protocol | Hazard rate | 0.95 (0.02) |
| | (censoring cases before $\alpha$) | Cumulative incidence | 0.93 (0.04) |
| | | Incidence rate | 0.95 (0.02) |

Table 1: Results of the data analysis performed on the Pfizer dataset. Vaccine efficacy estimates are shown for day 112.

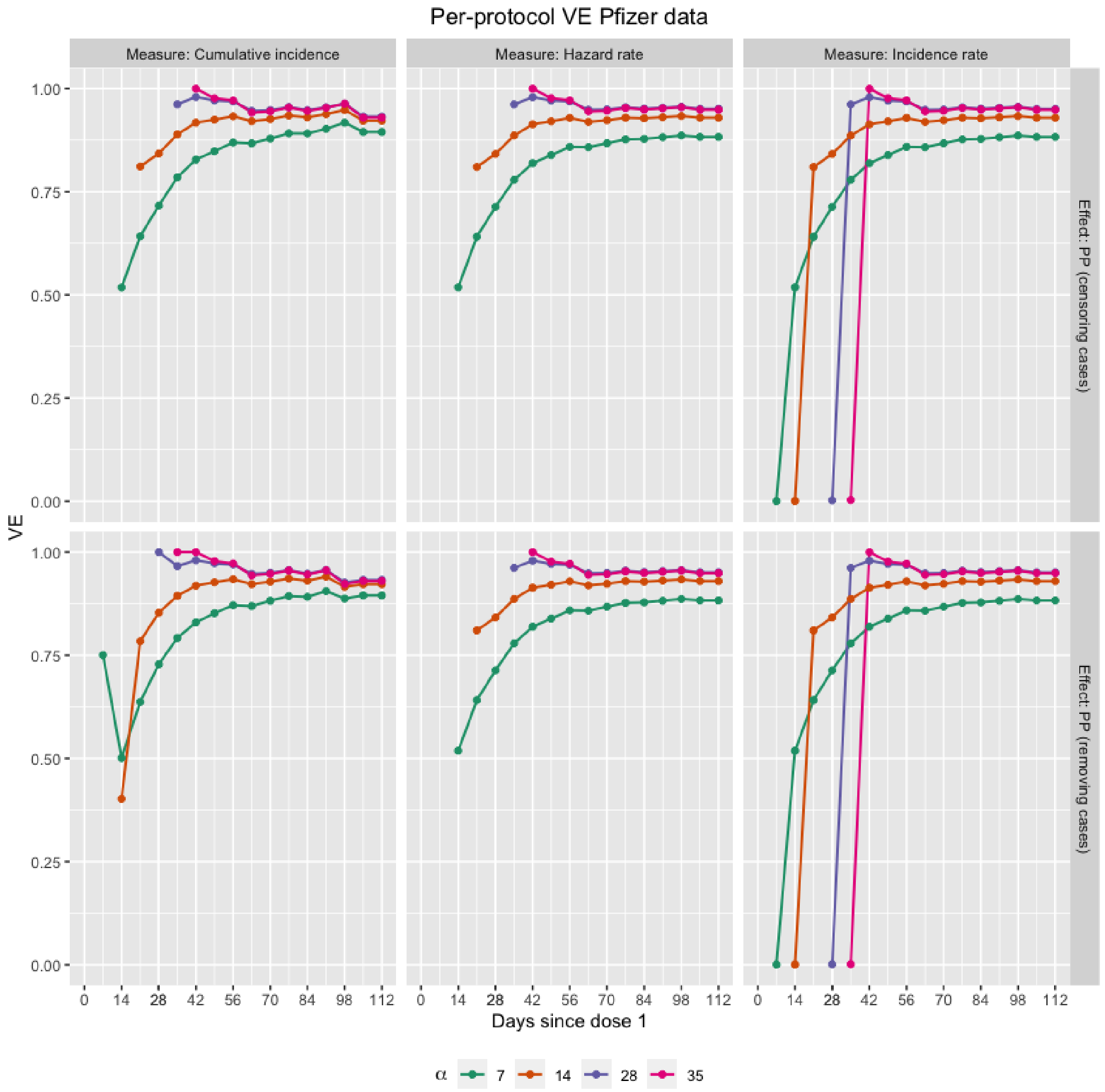


Figure 10: Per-protocol results of the data analysis performed on the Pfizer dataset, investigating different lengths of the ramp-up period ($\alpha = 7, 14, 28, 35$ days). Vaccine efficacy estimates are shown over time, for the cumulative incidence, hazard rate and incidence rate as risk measure.

| **Ramp-up period** | **Effect** | **Risk measure** | **VE (SE)** |
|---|---|---|---|
| | Intention-to-treat | Hazard rate | 0.55 (0.04) |
| | | Cumulative incidence | 0.54 (0.10) |
| | | Incidence rate | 0.55 (0.04) |
| $\boldsymbol{\alpha}$ **= 7 days** | Per-protocol | Hazard rate | 0.58 (0.04) |
| cases before $\alpha$: | (removing cases before $\alpha$) | Cumulative incidence | 0.56 (0.10) |
| 4 % of placebo cases | | Incidence rate | 0.58 (0.04) |
| 10 % of vaccine cases | Per-protocol | Hazard rate | 0.58 (0.04) |
| | (censoring cases before $\alpha$) | Cumulative incidence | 0.56 (0.10) |
| | | Incidence rate | 0.58 (0.04) |
| $\boldsymbol{\alpha}$ **= 14 days** | Per-protocol | Hazard rate | 0.67 (0.04) |
| cases before $\alpha$: | (removing cases before $\alpha$) | Cumulative incidence | 0.61 (0.11) |
| 18 % of placebo cases | | Incidence rate | 0.67 (0.04) |
| 39 % of vaccine cases | Per-protocol | Hazard rate | 0.67 (0.03) |
| | (censoring cases before $\alpha$) | Cumulative incidence | 0.61 (0.10) |
| | | Incidence rate | 0.67 (0.04) |
| $\boldsymbol{\alpha}$ **= 28 days** | Per-protocol | Hazard rate | 0.66 (0.05) |
| cases before $\alpha$: | (removing cases before $\alpha$) | Cumulative incidence | 0.58 (0.16) |
| 55 % of placebo cases | | Incidence rate | 0.66 (0.05) |
| 65 % of vaccine cases | Per-protocol | Hazard rate | 0.66 (0.05) |
| | (censoring cases before $\alpha$) | Cumulative incidence | 0.58 (0.15) |
| | | Incidence rate | 0.66 (0.05) |
| $\boldsymbol{\alpha}$ **= 35 days** | Per-protocol | Hazard rate | 0.69 (0.06) |
| cases before $\alpha$: | (removing cases before $\alpha$) | Cumulative incidence | 0.58 (0.18) |
| 69 % of placebo cases | | Incidence rate | 0.69 (0.06) |
| 78 % of vaccine cases | Per-protocol | Hazard rate | 0.69 (0.06) |
| | (censoring cases before $\alpha$) | Cumulative incidence | 0.58 (0.18) |
| | | Incidence rate | 0.69 (0.06) |

Table 2: Results of the data analysis performed on the Janssen dataset. Vaccine efficacy estimates are shown for day 125.

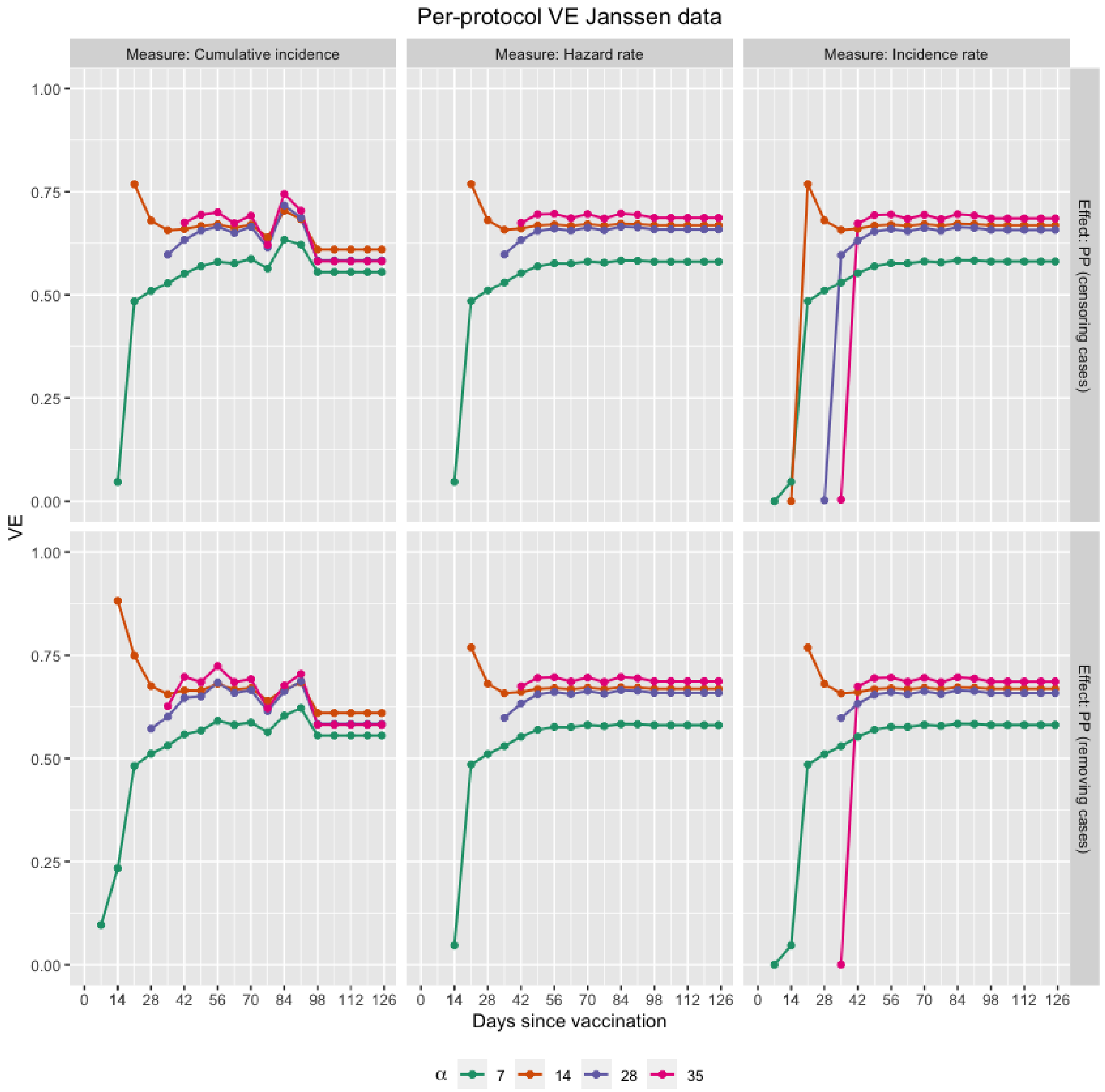


Figure 11: Per-protocol results of the data analysis performed on the Janssen dataset, investigating different lengths of the ramp-up period ($\alpha$ = 7, 14, 28, 35 days). Vaccine efficacy estimates are shown over time, for the cumulative incidence, hazard rate and incidence rate as risk measure.

##### C.4.3 Vaccine efficacy if ramp-up period can be eliminated

**Methods**

For both trials, the second hypothetical estimand (section 3.2.2) was estimated every week using model (12), where either the 3 model parameters were estimated or 1 or 2 parameters were specified, as described in section C.3.4. The expressions (13), (15) or (17) for the initial parameter values were minimized using the `gridSearch` function in R. These initial values were updated by minimizing the sum of differences in restricted mean survival times (14), (16) and (18), using the `optim` function. The empirical restricted mean survival times were obtained using the `rmst2` function of the `survRM2` package [30]. In addition, penalties were added to the expressions (14), (16) and (18) to make sure that the parameter estimates were bounded: $\hat{\psi} \geq 0$, $\hat{\rho} \in [0, 1]$ and $\hat{\alpha} \in [0, L/2]$. Details about the implementation in R are given in Appendix D.2.4.

For methods 1 and 2 (section C.3.4) the length of the ramp-up period $\alpha$ needs to be specified. The same lengths as in the study protocols, i.e. $\alpha = 14$ for the Janssen trial and $\alpha = 28$ for the Pfizer trial, were used. In addition, a shorter period, i.e. $\alpha = 14$ days, was also investigated for the Pfizer trial since from the Kaplan-Meier plot (figure 8) it looked like the vaccine had already an effect from that visit. For method 1 (section C.3.4), the parameter $\rho$ that indicates how effective the vaccine is during the ramp-up time, needs also be specified. For the Janssen trial, $\rho = 0$ was chosen as the survival curves completely coincide during the first 2 weeks (figure 9). In case of the Pfizer trial, the value $\rho = 0.40$ was specified if $\alpha = 28$ days and $\rho = 0.10$ if $\alpha = 14$ days. These values for $\rho$ were chosen as an example since the vaccine has a very limited effect during the first 14 days and a larger, but limited effect during the first 28 days.

The hypothetical estimand was estimated using expression (19), where the survival curve in the placebo arm was estimated using the Kaplan-Meier estimator. Standard errors (SE) were obtained using 1000 non-parametric bootstrap replications [8]. However, because of the computational complexity, SEs were only calculated for parameter $\psi$ and the hypothetical vaccine efficacy using method 1.

**Results**

Results for the data analysis of the vaccine efficacy if the ramp-up period can be eliminated on the Janssen and Pfizer dataset are given in tables 3 and 4 and figures 12 - 18 below. For the Pfizer trial, methods 1 and 2 with $\alpha = 28$ days return parameter estimates for which the mapped survival function (12) coincides well with the observed vaccine survival curve (figure 13). Moreover, the other methods with shorter ramp-up times ($\alpha = 14$ or $\hat{\alpha} = 9.57$) lead to survival curves that fit the data almost perfectly (figures 14 and 15). The counterfactual survival curves 'as if there was no ramp-up time' are also shown on these plots. Using these survival curves, the hypothetical vaccine efficacy estimates are calculated, as shown in table 3 and figure 12. All estimates are close to each other and the per-protocol estimators.

For the Janssen trial, methods 1 and 2 lead to mapped survival functions that fit the observed curve fairly well (figure 17), although the increase in cumulative incidence is a bit too big after the ramp-up period. As a result, the hypothetical vaccine efficacy estimates drop significantly over time (figure 16). Method 3, where all 3 parameters are estimated, does not lead to a good approximation of the observed survival curve (figure

18).

Pfizer COVID-19 trial

| Method | Parameters (SE) | | | Hypothetical VE (SE) |
|---|---|---|---|---|
| **Method 1** | | | | |
| $\psi$: estimated<br>$\rho$ and $\alpha$: specified | $\rho = 0.40$ | $\hat{\psi} = 2.55$ (0.44) | $\alpha = 28.00$ | 0.93 (0.04) |
| **Method 2** | | | | |
| $\rho$ and $\psi$: estimated<br>$\alpha$: specified | $\hat{\rho} = 0.33$ | $\hat{\psi} = 2.56$ | $\alpha = 28.00$ | 0.93 |
| **Method 3** | | | | |
| $\rho$, $\psi$ and $\alpha$: estimated | $\hat{\rho} = 0.00$ | $\hat{\psi} = 2.69$ | $\hat{\alpha} = 9.57$ | 0.95 |
| **Method 1** | | | | |
| $\psi$: estimated<br>$\rho$ and $\alpha$: specified | $\rho = 0.10$ | $\hat{\psi} = 2.84$ | $\alpha = 14.00$ | 0.95 |
| **Method 2** | | | | |
| $\rho$ and $\psi$: estimated<br>$\alpha$: specified | $\hat{\rho} = 0.24$ | $\hat{\psi} = 2.51$ | $\alpha = 14.00$ | 0.93 |

Table 3: Results of the hypothetical vaccine efficacy estimates 'if there was no ramp-up time' on the Pfizer dataset. Vaccine efficacy estimates are shown for day 112.

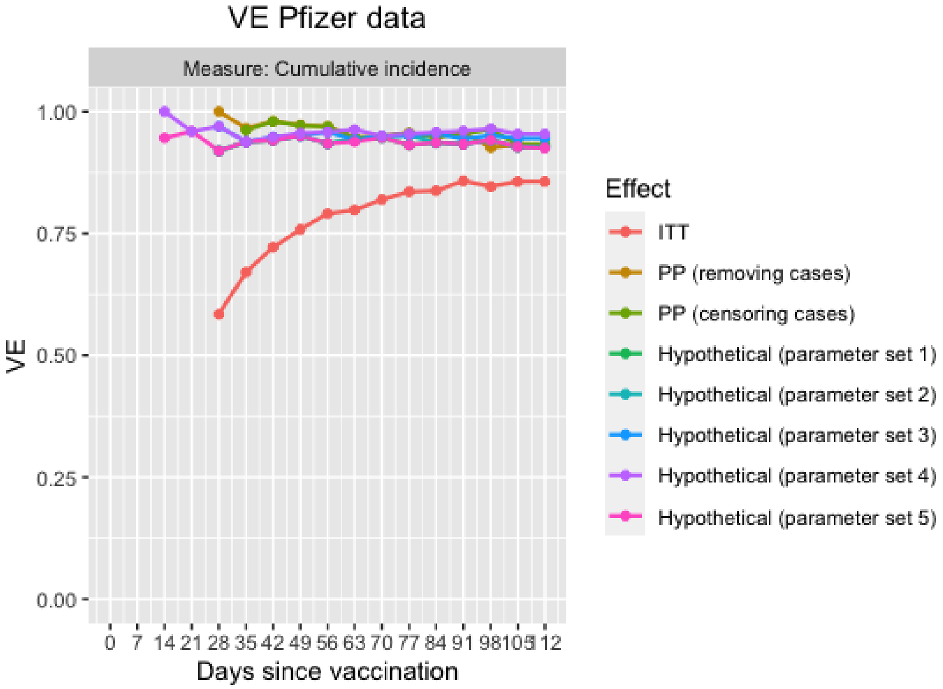


Figure 12: Results of the data analysis performed on the Pfizer dataset. Vaccine efficacy estimates are shown over time for the cumulative incidence as risk measure. Every point represents the VE estimate that would be obtained if the trial was stopped at the corresponding visit and all information up till that visit was used.
Parameter set 1: $\rho = 0.40$, $\hat{\psi} = 2.55$ and $\alpha = 28.00$
Parameter set 2: $\hat{\rho} = 0.33$, $\hat{\psi} = 2.56$ and $\alpha = 28.00$
Parameter set 3: $\hat{\rho} = 0.00$, $\hat{\psi} = 2.69$ and $\alpha = 9.57$
Parameter set 4: $\rho = 0.10$, $\hat{\psi} = 2.84$ and $\alpha = 14.00$
Parameter set 5: $\hat{\rho} = 0.24$, $\hat{\psi} = 2.51$ and $\alpha = 14.00$

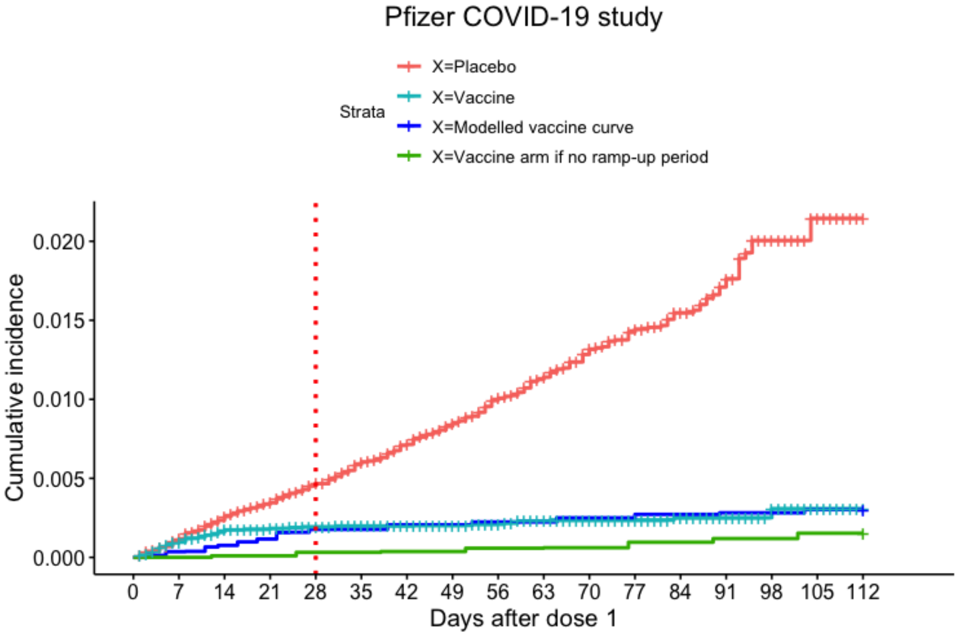


(a) Method 1: $\rho = 0.40$, $\hat{\psi} = 2.55$ and $\alpha = 28$

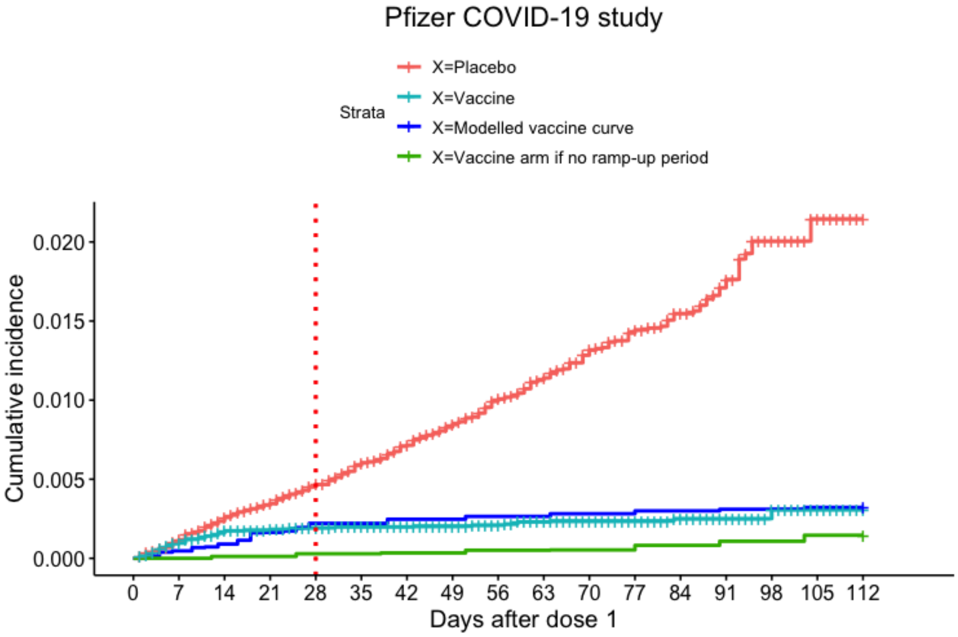


(b) Method 2: $\hat{\rho} = 0.33$, $\hat{\psi} = 2.56$ and $\alpha = 28$

Figure 13: Data of the Pfizer COVID-19 vaccine trial, together with the modeled vaccine survival curve (model (12)) and the counterfactual vaccine survival curve if there was no ramp-up time, for different parameter values.

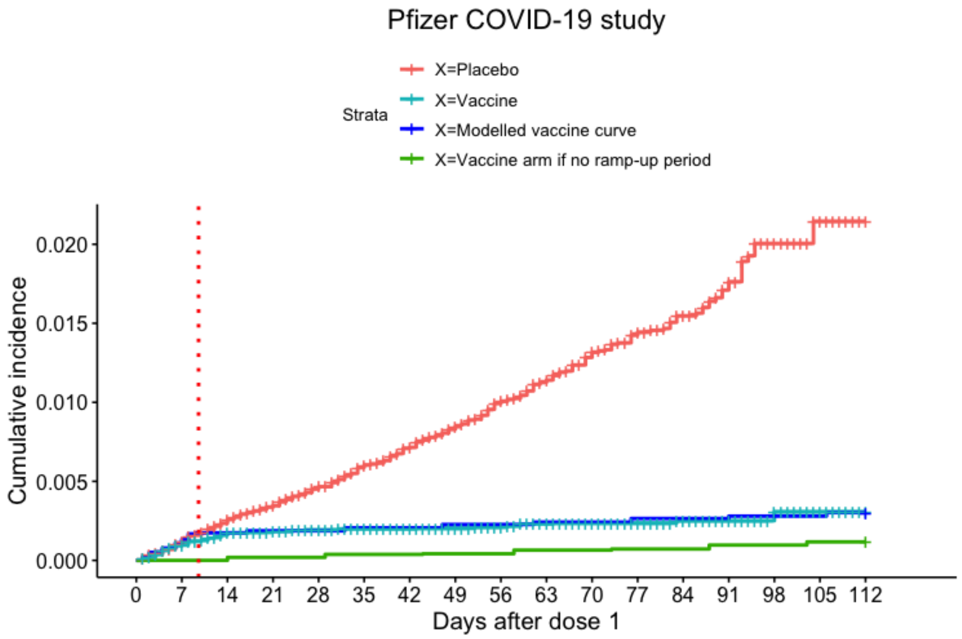


(a) Method 3: $\hat{\rho} = 0.00$, $\hat{\psi} = 2.69$ and $\hat{\alpha} = 9.57$

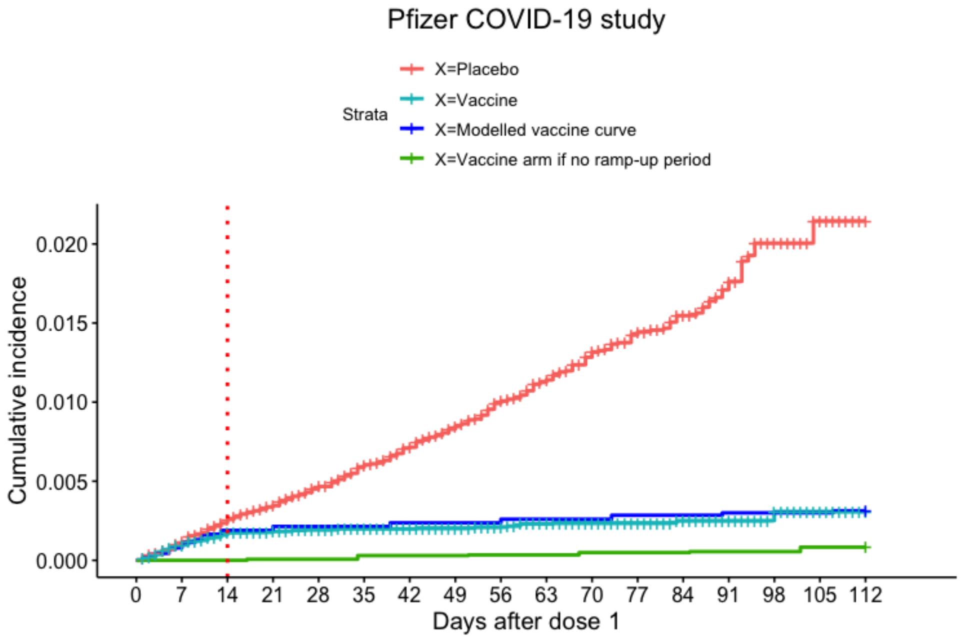


(b) Method 1: $\rho = 0.10$, $\hat{\psi} = 2.84$ and $\alpha = 14$

Figure 14: Data of the Pfizer COVID-19 vaccine trial, together with the modelled vaccine survival curve (model (12)) and the counterfactual vaccine survival curve if there was no ramp-up time, for different parameter values.

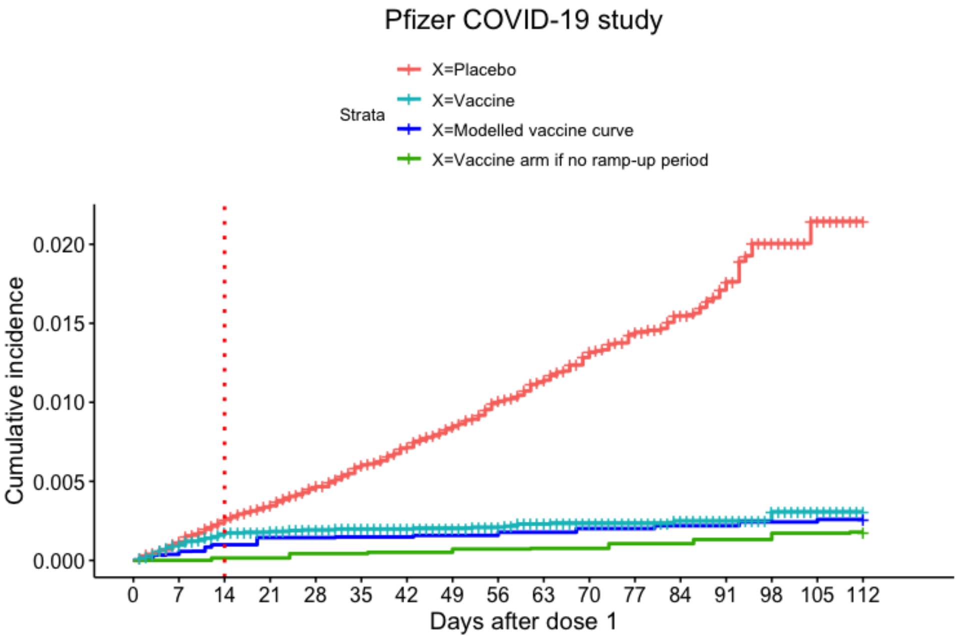


(a) Method 2: $\hat{\rho} = 0.24$, $\hat{\psi} = 2.51$ and $\alpha = 14$

Figure 15: Data of the Pfizer COVID-19 vaccine trial, together with the modeled vaccine survival curve (model (12)) and the counterfactual vaccine survival curve if there was no ramp-up time, for different parameter values.

Janssen COVID-19 trial

| **Method** | **Parameters (SE)** | | | **Hypothetical VE (SE)** |
|---|---|---|---|---|
| **Method 1** | | | | |
| $\psi$: estimated<br>$\rho$ and $\alpha$: specified | $\rho = 0.00$ | $\hat{\psi} = 1.58$ (0.35) | $\alpha = 14.00$ | 0.66 (0.13) |
| **Method 2** | | | | |
| $\rho$ and $\psi$: estimated<br>$\alpha$: specified | $\hat{\rho} = 0.07$ | $\hat{\psi} = 1.59$ | $\alpha = 14.00$ | 0.66 |
| **Method 3** | | | | |
| $\rho$, $\psi$ and $\alpha$: estimated | $\hat{\rho} = 0.01$ | $\hat{\psi} = 2.09$ | $\hat{\alpha} = 19.24$ | 0.84 |

Table 4: Results of the hypothetical vaccine efficacy estimates 'if there was no ramp-up time' on the Janssen dataset. Vaccine efficacy estimates are shown for day 125.

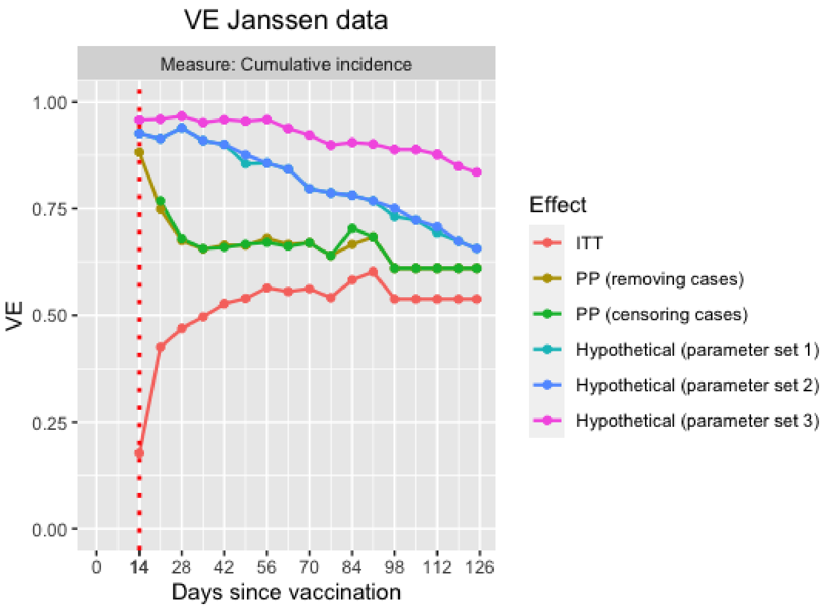


Figure 16: Results of the data analysis performed on the Janssen dataset. Vaccine efficacy estimates are shown over time for the cumulative incidence as risk measure. Every point represents the VE estimate that would be obtained if the trial was stopped at the corresponding visit and all information up till that visit was used.
Parameter set 1: $\rho = 0.00$, $\hat{\psi} = 1.58$ and $\alpha = 14.00$
Parameter set 2: $\hat{\rho} = 0.07$, $\hat{\psi} = 1.59$ and $\alpha = 14.00$
Parameter set 3: $\hat{\rho} = 0.01$, $\hat{\psi} = 2.09$ and $\hat{\alpha} = 19.24$

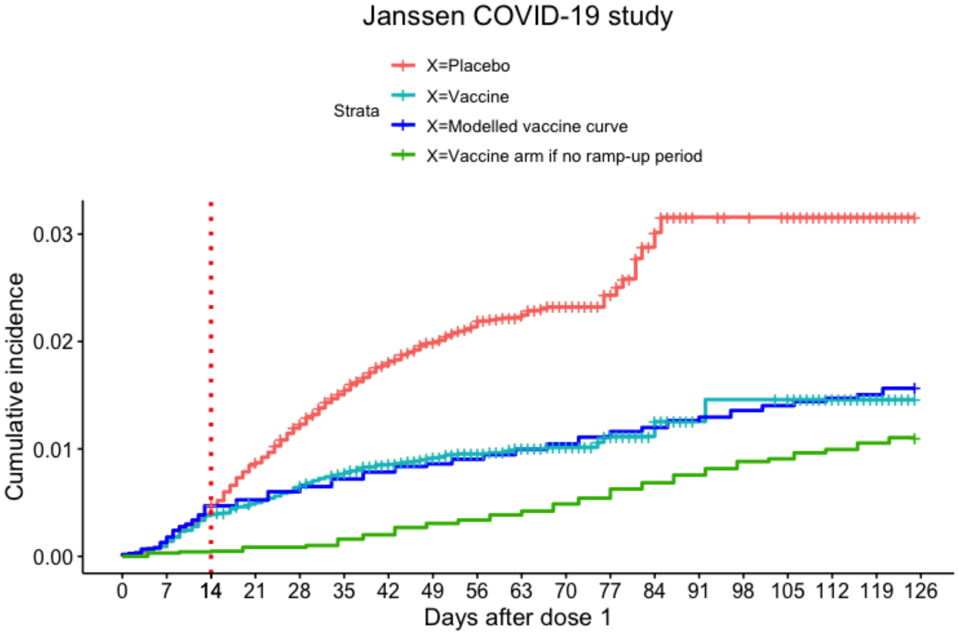


(a) Method 1: $\rho = 0.00$, $\hat{\psi} = 1.58$ and $\alpha = 14$

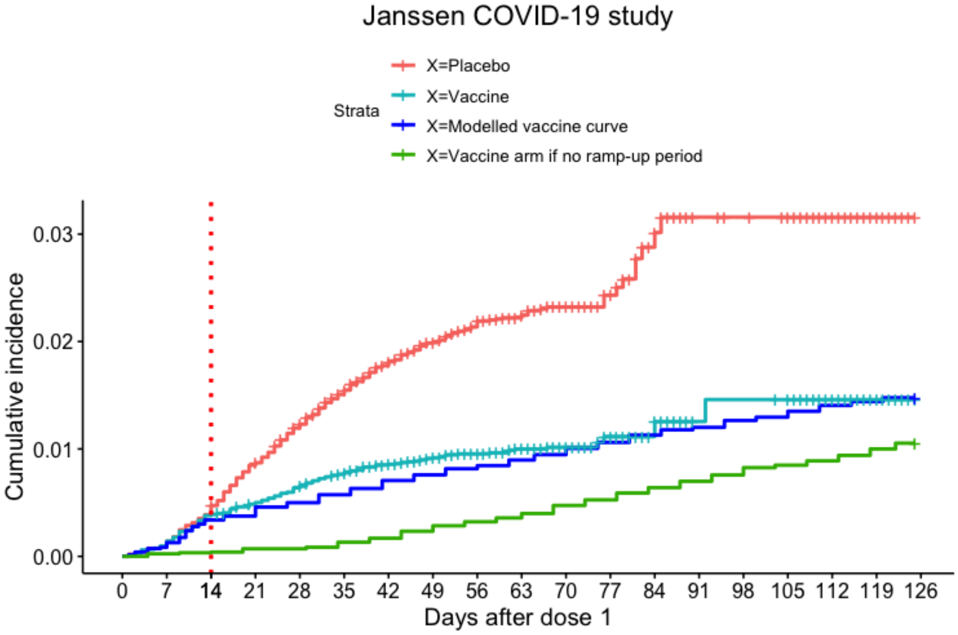


(b) Method 2: $\hat{\rho} = 0.07$, $\hat{\psi} = 1.59$ and $\alpha = 14$

Figure 17: Data of the Janssen COVID-19 vaccine trial, together with the modeled vaccine survival curve (model (12)) and the counterfactual vaccine survival curve if there was no ramp-up time, for different parameter values.

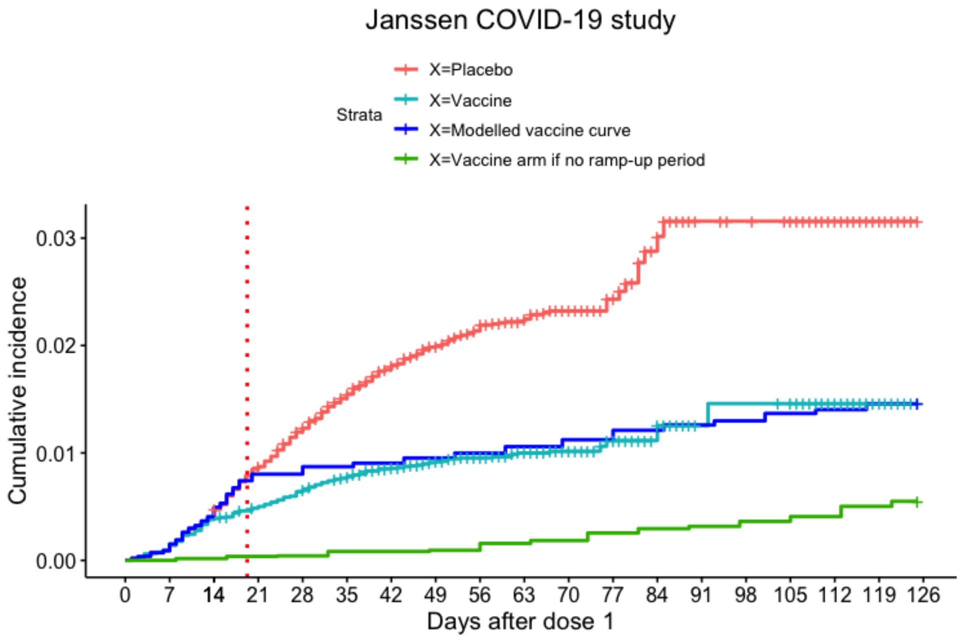


(a) Method 3: $\hat{\rho} = 0.01$, $\hat{\psi} = 2.09$ and $\hat{\alpha} = 19.24$

Figure 18: Data of the Janssen COVID-19 vaccine trial, together with the modeled vaccine survival curve (model (12)) and the counterfactual vaccine survival curve if there was no ramp-up time, for different parameter values.

### D R-code

In this section, we provide R-code for construction of the Pfizer and Janssen datasets and to implement the discussed vaccine efficacy estimators. R-files with this code can be found in the online supplementary files.

#### D.1 Construction of the datasets

##### D.1.1 Pfizer dataset

First, we load some packages:

```
library(ggplot2)
library(survival)
library(survminer)
```

Next, we set a seed to make the code reproducible:

```
set.seed(1)
```

Next, we make two vectors with the number of patients at risk per 7 days, separately for the vaccine and the placebo arm. These numbers are based on Figure 2 in [12].

```
#Number of patients at risk in the vaccine arm
at.risk.V = c(21314,21230,21054,20481,19314,18377,17702,17186,15464,
14038,12169,9591,6403,3374,1463,398,0)
#Number of patients at risk in the placebo arm
at.risk.PB = c(21258,21170,20970,20366,19209,18218,17578,17025,15290,
13876,11994,9471,6294,3301,1449,398,0)
```

Now we make two vectors with the cumulative number of cases per 7 days, separately for the vaccine and the placebo arm. These numbers are again based on Figure 2 in [12].

```
#Cumulative number of cases in the vaccine arm
cum.cases.V = c(0,21,37,39,41,42,42,43,44,47,48,48,49,49,50,50,50)
#Cumulative number of cases in the placebo arm
cum.cases.PB = c(0,25,55,73,97,123,143,166,192,212,235,249,257,267,274,275,275)
```

Next, we make two vectors with the observed number of cases per week, since we will work with these numbers instead of the cumulative numbers:

```
cases.V = c() #Number of cases in the vaccine arm
cases.PB = c() #Number of cases in the placebo arm
cases.V[1] = 0
cases.PB[1] = 0
```

```
for(i in 2:length(cum.cases.PB)){
cases.V[i] = cum.cases.V[i]-cum.cases.V[i-1]
cases.PB[i] = cum.cases.PB[i]-cum.cases.PB[i-1]
}
cases.V #Number of cases in the vaccine arm (per 7 days)
cases.PB #Number of cases in the placebo arm (per 7 days)
```

Next, we calculate the number of censored patients per 7 days by subtracting the cases from the patients at risk. We put these numbers again in a vector per treatment arm:

```
censored.V = c() #Number of censored patients in the vaccine arm
censored.PB = c() #Number of censored patients in the placebo arm
censored.V[1] = 0
censored.PB[1] = 0
for(i in 2:length(cum.cases.PB)){
censored.V[i] = at.risk.V[i-1] - at.risk.V[i] - cases.V[i]
censored.PB[i] = at.risk.PB[i-1] - at.risk.PB[i] - cases.PB[i]
}
censored.V #Number of censored patients in the vaccine arm (per 7 days)
censored.PB #Number of censored patients in the placebo arm (per 7 days)
```

Now we can construct the Pfizer dataset. First, we specify the study visits (every 7 days starting from baseline to day 112) and the length of the ramp-up period:

```
visits = seq(0,112,by=7)  #Study visits in the Pfizer trial
censor.day = 112 #End of the trial: patients who are still in the trial
at this day are administratively censored.
alpha = 28 #Ramp-up period, starting from randomization/first dose
```

Next, we make vectors with an identification number per patient, the patients at risk, a treatment arm indicator, the follow-up times and censoring indicators, that will be used to construct the placebo dataset. These vectors will be updated in the next steps.

```
#Follow-up number for the placebo patients
PB.patients = 1:at.risk.PB[1]
#Number of placebo patients at risk at randomization
PB.patients.at.risk = PB.patients
#Placebo arm is indicated with X=0
PB.X = rep(0,times = at.risk.PB[1])
#The vector ‘PB.status’ will be used to indicate whether a patient is a case (1),
or is censored (0)
PB.status = rep(0,times = at.risk.PB[1]) #At the start, all statuses are set to 0
#The vector ‘PB.time’ will be used to indicate a patient’s failure/censoring time
PB.time = rep(112,times = at.risk.PB[1]) #Everybody’s follow-up time is set
at the end of the trial
```

Now we run a for-loop over all visits. In every step, we consider the total number of cases and censored patients for that visit.

```
for(i in 2:length(visits)){
t.start = visits[i-1]+1 #Start of this time interval
t.end = visits[i] #End of this time interval

#Cases in this interval
cases = cases.PB[i] #Number of cases in this interval
#Cases in this interval are randomly chosen from the patients who are still at risk
cases.patients = sample(PB.patients.at.risk,size=cases)
#Infection times are chosen randomly in this interval for these patients
cases.time = sample(t.start:t.end,size=cases,replace=TRUE)
PB.time[cases.patients] = cases.time #The ‘PB.time’ vector is updated
PB.status[cases.patients] = 1 #For these cases, the infection status is set to 1
#Cases are removed from the at risk set
PB.patients.at.risk = setdiff(PB.patients.at.risk, cases.patients)

#Censored patients in this interval
censored = censored.PB[i] #Number of censored patients in this interval
#Censored patients in this interval are randomly chosen from the patients
who are still at risk
censored.patients = sample(PB.patients.at.risk,size=censored)
#Censoring times are chosen randomly in this interval for these patients
censored.time = sample(t.start:t.end,size=censored,replace=TRUE)
PB.time[censored.patients] = censored.time #The ‘PB.time’ vector is updated
#For these censored patients, the infection status is set to 0
PB.status[censored.patients] = 0
#Censored patients are removed from the at risk set
PB.patients.at.risk = setdiff(PB.patients.at.risk, censored.patients)
}
```

Finally, we construct the placebo dataset:

```
PB.data = data.frame(PB.patients,PB.X,PB.time,PB.status,stringsAsFactors = FALSE)
colnames(PB.data) = c("patient","X","time","status")
head(PB.data)
```

Next, we create the dataset of the patients in the vaccine arm in a similar way:

```
#Follow-up number for the vaccine patients
V.patients = 1:at.risk.V[1]
#Number of vaccine patients at risk at randomization
```

```
V.patients.at.risk = V.patients
#Vaccine arm is indicated with X=1
V.X = rep(0,times = at.risk.V[1])
#The vector ‘V.status’ will be used to indicate whether a patient is a case (1),
or is censored (0)
V.status = rep(0,times = at.risk.V[1]) #At the start, all statuses are set to 0
#The vector ‘V.time’ will be used to indicate a patient’s failure/censoring time
V.time = rep(112,times = at.risk.V[1]) #Everybody’s follow-up time is set
at the end of the trial

for(i in 2:length(visits)){
t.start = visits[i-1]+1 #Start of this time interval
t.end = visits[i] #End of this time interval

#Cases in this interval
cases = cases.V[i] #Number of cases in this interval
#Cases in this interval are randomly chosen from the patients who are still at risk
cases.patients = sample(V.patients.at.risk,size=cases)
#Infection times are chosen randomly in this interval for these patients
cases.time = sample(t.start:t.end,size=cases,replace=TRUE)
V.time[cases.patients] = cases.time #The ‘V.time’ vector is updated
V.status[cases.patients] = 1 #For these cases, the infection status is set to 1
#Cases are removed from the at risk set
V.patients.at.risk = setdiff(V.patients.at.risk, cases.patients)

#Censored patients in this interval
censored = censored.V[i] #Number of censored patients in this interval
#Censored patients in this interval are randomly chosen from the patients
who are still at risk
censored.patients = sample(V.patients.at.risk,size=censored)
#Censoring times are chosen randomly in this interval for these patients
censored.time = sample(t.start:t.end,size=censored,replace=TRUE)
V.time[censored.patients] = censored.time #The ‘V.time’ vector is updated
#For these censored patients, the infection status is set to 0
V.status[censored.patients] = 0
#Censored patients are removed from the at risk set
V.patients.at.risk = setdiff(V.patients.at.risk, censored.patients)
}
```

Next, we construct the vaccine dataset:

```
V.data = data.frame(V.patients,V.X,V.time,V.status,stringsAsFactors = FALSE)
```

```
colnames(V.data) = c("patient","X","time","status")
head(V.data)
```

Finally, the total dataset can be created by combining the placebo and vaccine dataset:

```
data = rbind(PB.data,V.data)
data.pfizer = data
head(data)
```

The dataset can be saved in a txt-file:

```
write.table(data.pfizer, file = "data.pfizer.txt", sep = "\t",
row.names = TRUE, col.names = TRUE)
```

A Kaplan-Meier curve can be created to get insight into the dataset:

```
data.plot = data
data.plot$X = ifelse(data.plot$X==0,yes="Placebo",no="Vaccine")

p = ggsurvplot(survfit(Surv(time, status) ~ X, data = data.plot),
title = "Pfizer COVID-19 study", risk.table = TRUE,cumevents = TRUE,
break.time.by=7, risk.table.height = 0.15, cumevents.height = 0.15,
risk.table.y.text = FALSE, cumevents.y.text = FALSE, fontsize = 3,
risk.table.title = "No. at Risk", cumevents.title = "Cumulative No. of Events",
xlab = "Days after dose 1", ylab = "Cumulative incidence",
xlim=c(0,119),ylim=c(0,0.024),size=1,fun="event")

p$plot = p$plot + theme(plot.title = element_text(hjust = 0.5))
p$plot = p$plot + scale_x_continuous(breaks=c(seq(0,119,7),alpha))
p$plot = p$plot+ geom_vline(xintercept = alpha, linetype="dotted",
              color = "pink", size=1)
p$plot = p$plot+ geom_vline(xintercept = 0, linetype="dotted",
              color = "grey", size=1)
p$plot = p$plot+ geom_vline(xintercept = 21, linetype="dotted",
              color = "grey", size=1)
p$table = p$table + theme_cleantable()
p$table = p$table + theme(plot.title = element_text(size = 12))
p$cumevents = p$cumevents + theme_cleantable()
p$cumevents = p$cumevents + theme(plot.title = element_text(size = 12))
p
```

##### D.1.2 Janssen dataset

First, we load some packages:

```
library(ggplot2)
library(survival)
library(survminer)
```

Next, we set a seed to make the code reproducible:

```
set.seed(1)
```

Next, we make two vectors with the number of patients at risk per 7 days, separately for the vaccine and the placebo arm. These numbers are based on Figure 1 in [13].

```
#Number of patients at risk in the vaccine arm
at.risk.V =c(19744,19725,19669,19642,19612,19578,18541,14909,10930,7831,
3998,1468,713,484,483,482,142,31,0)
#Number of patients at risk in the placebo arm
at.risk.PB =  c(19822,19804,19745,19652,19579,19488,18411,14814,10823,
7740,3876,1439,708,485,482,480,133,27,0)
```

Now we make two vectors with the cumulative number of cases per 7 days, separately for the vaccine and the placebo arm. These numbers are again based on Figure 1 in [13].

```
#Cumulative number of cases in the vaccine arm
cum.cases.V = c(0,19,75,96,126,151,168,178,184,188,189,191,191,192,193,
193,193,193,193)
#Cumulative number of cases in the placebo arm
cum.cases.PB = c(0,18,77,168,237,299,351,387,407,416,423,425,430,432,
432,432,432,432,432)
```

Next, we make two vectors with the number of cases observed per week, since we will work with these numbers instead of the cumulative numbers:

```
cases.V = c() #Number of cases in the vaccine arm
cases.PB = c() #Number of cases in the placebo arm
cases.V[1] = 0
cases.PB[1] = 0
for(i in 2:length(cum.cases.PB)){
cases.V[i] = cum.cases.V[i]-cum.cases.V[i-1]
cases.PB[i] = cum.cases.PB[i]-cum.cases.PB[i-1]
}
cases.V #Number of cases in the vaccine arm (per 7 days)
cases.PB #Number of cases in the placebo arm (per 7 days)
```

Next, we calculate the number of censored patients per 7 days by subtracting the cases from the patients at risk. We put these numbers again in a vector per treatment arm:

```
censored.V = c() #Number of censored patients in the vaccine arm
censored.PB = c() #Number of censored patients in the placebo arm
censored.V[1] = 0
censored.PB[1] = 0
for(i in 2:length(cum.cases.PB)){
censored.V[i] = at.risk.V[i-1] - at.risk.V[i] - cases.V[i]
censored.PB[i] = at.risk.PB[i-1] - at.risk.PB[i] - cases.PB[i]
}
censored.V #Number of censored patients in the vaccine arm (per 7 days)
censored.PB #Number of censored patients in the placebo arm (per 7 days)
```

Now we can construct the Janssen dataset. First, we specify the study visits (every 7 days starting from baseline to day 126) and the length of the ramp-up period:

```
visits = seq(0,126,by= 7) #Study visits in the Janssen trial
censor.day = 126 #End of the trial: patients who are still in the trial
at this day are administratively censored.
alpha = 14 #Ramp-up period, starting from randomization/vaccination
```

Next, we make vectors with follow-up numbers per patient, the patients at risk, a treatment arm indicator, the follow-up times and censoring indicators, that will be used to construct the placebo dataset. These vectors will be updated in the next steps.

```
#Follow-up number for the placebo patients
PB.patients = 1:at.risk.PB[1]
#Number of placebo patients at risk at randomization
PB.patients.at.risk = PB.patients
#Placebo arm is indicated with X=0
PB.X = rep(0,times = at.risk.PB[1])
#The vector ‘PB.status’ will be used to indicate whether a patient is a case (1),
or is censored (0)
PB.status = rep(0,times = at.risk.PB[1]) #At the start, all statuses are set to 0
#The vector ‘PB.time’ will be used to indicate a patient’s failure/censoring time
PB.time = rep(126,times = at.risk.PB[1]) #Everybody’s follow-up time is set
at the end of the trial
```

Now we run a for-loop over all visits. In every step, we consider the total number of cases and censored patients for that visit.

```
for(i in 2:length(visits)){
t.start = max(visits[i-1],1) #Start of this time interval
t.end = visits[i]-1 #End of this time interval
```

```
#Cases in this interval
cases = cases.PB[i] #Number of cases in this interval
#Cases in this interval are randomly chosen from the patients who are still at risk
cases.patients = sample(PB.patients.at.risk,size=cases)
#Infection times are chosen randomly in this interval for these patients
cases.time = sample(t.start:t.end,size=cases,replace=TRUE)
PB.time[cases.patients] = cases.time #The ‘PB.time’ vector is updated
PB.status[cases.patients] = 1 #For these cases, the infection status is set to 1
#Cases are removed from the at risk set
PB.patients.at.risk = setdiff(PB.patients.at.risk, cases.patients)

#Censored patients in this interval
censored = censored.PB[i] #Number of censored patients in this interval
#Censored patients in this interval are randomly chosen from the patients
who are still at risk
censored.patients = sample(PB.patients.at.risk,size=censored)
#Censoring times are chosen randomly in this interval for these patients
censored.time = sample(t.start:t.end,size=censored,replace=TRUE)
PB.time[censored.patients] = censored.time #The ‘PB.time’ vector is updated
#For these censored patients, the infection status is set to 0
PB.status[censored.patients] = 0
#Censored patients are removed from the at risk set
PB.patients.at.risk = setdiff(PB.patients.at.risk, censored.patients)
}
```

Finally, we construct the placebo dataset:

```
PB.data = data.frame(PB.patients,PB.X,PB.time,PB.status,stringsAsFactors = FALSE)
colnames(PB.data) = c("patient","X","time","status")
head(PB.data)
```

Next, we create the dataset of the patients in the vaccine arm in a similar way:

```
#Follow-up number for the vaccine patients
V.patients = 1:at.risk.V[1]
#Number of vaccine patients at risk at randomization
V.patients.at.risk = V.patients
#Vaccine arm is indicated with X=1
V.X = rep(0,times = at.risk.V[1])
#The vector ‘V.status’ will be used to indicate whether a patient is a case (1),
or is censored (0)
V.status = rep(0,times = at.risk.V[1]) #At the start, all statuses are set to 0
#The vector ‘V.time’ will be used to indicate a patient’s failure/censoring time
```

```
V.time = rep(112,times = at.risk.V[1]) #Everybody’s follow-up time is set
at the end of the trial

for(i in 2:length(visits)){
t.start = max(visits[i-1],1) #Start of this time interval
t.end = visits[i]-1 #End of this time interval

#Cases in this interval
cases = cases.V[i] #Number of cases in this interval
#Cases in this interval are randomly chosen from the patients who are still at risk
cases.patients = sample(V.patients.at.risk,size=cases)
#Infection times are chosen randomly in this interval for these patients
cases.time = sample(t.start:t.end,size=cases,replace=TRUE)
V.time[cases.patients] = cases.time #The ‘V.time’ vector is updated
V.status[cases.patients] = 1 #For these cases, the infection status is set to 1
#Cases are removed from the at risk set
V.patients.at.risk = setdiff(V.patients.at.risk, cases.patients)

#Censored patients in this interval
censored = censored.V[i] #Number of censored patients in this interval
#Censored patients in this interval are randomly chosen from the patients
who are still at risk
censored.patients = sample(V.patients.at.risk,size=censored)
#Censoring times are chosen randomly in this interval for these patients
censored.time = sample(t.start:t.end,size=censored,replace=TRUE)
V.time[censored.patients] = censored.time #The ‘V.time’ vector is updated
#For these censored patients, the infection status is set to 0
V.status[censored.patients] = 0
#Censored patients are removed from the at risk set
V.patients.at.risk = setdiff(V.patients.at.risk, censored.patients)
}
```

Next, we construct the vaccine dataset:

```
V.data = data.frame(V.patients,V.X,V.time,V.status,stringsAsFactors = FALSE)
colnames(V.data) = c("patient","X","time","status")
head(V.data)
```

Finally, the total dataset can be created by combining the placebo and vaccine dataset:

```
data = rbind(PB.data,V.data)
data.jnj = data
head(data)
```

The dataset can be saved in a txt-file:

```
write.table(data.jnj, file = "data.jnj.txt", sep = "\t",
row.names = TRUE, col.names = TRUE)
```

A Kaplan-Meier curve can be created to get insight into the dataset:

```
data.plot = data
data.plot$X = ifelse(data.plot$X==0,yes="Placebo",no="Vaccine")

p = ggsurvplot(survfit(Surv(time, status) ~ X, data = data.plot),
title = "Janssen COVID-19 study",
risk.table = TRUE,cumevents = TRUE,   break.time.by=7,
risk.table.height = 0.15, cumevents.height = 0.15,
risk.table.y.text = FALSE,
cumevents.y.text = FALSE,
fontsize = 3,
risk.table.title = "No. at Risk",
cumevents.title = "Cumulative No. of Events",
xlab = "Days after dose 1",
ylab = "Cumulative incidence",xlim=c(0,126),ylim=c(0,0.035),size=1,fun="event")

p$plot = p$plot + theme(plot.title = element_text(hjust = 0.5))
p$plot = p$plot + scale_x_continuous(breaks=c(seq(0,126,7),alpha))
p$plot = p$plot+ geom_vline(xintercept = alpha, linetype="dotted",
                   color = "pink", size=1)
p$plot = p$plot+ geom_vline(xintercept = 0, linetype="dotted",
               color = "grey", size=1)
p$table <- p$table + theme_cleantable()
p$table <- p$table + theme(plot.title = element_text(size = 12))
p$cumevents <- p$cumevents + theme_cleantable()
p$cumevents <- p$cumevents + theme(plot.title = element_text(size = 12))
p
```

#### D.2 Vaccine efficacy estimators

First, we load some packages:

```
library(ggplot2)
library(survminer)
library(survival)
library(NMOF)
library(survRM2)
```

Next, we import a dataset and specify the length of the trial and the ramp-up period.

```
#Pfizer data
data = data.pfizer
censor.day = 112 #End of the trial
alpha = 28 #Ramp-up period

#Janssen data
data = data.jnj
censor.day = 125 #End of the trial
alpha = 14 #Ramp-up period
```

##### D.2.1 ITT effects

In this section, we show how the intention-to-treat effects can be estimated.

First, we specify the day at which we will estimate the vaccine efficacy, for example at the end of the trial:

```
t = censor.day
```

The ITT effect using the cumulative incidence as risk measure can be estimated by fitting a Kaplan-Meier curve:

```
data = data.jnj #Another dataset (e.g. data.pfizer) can also be used
#Kaplan-Meier survival curve
fit = survfit(Surv(time, status) ~ X, data = data)
#Cumulative incidence placebo arm
CI.PB= 1-summary(fit,time=c(t), extend = TRUE)$surv[1]
#Cumulative incidence vaccine arm
CI.V= 1-summary(fit,time=c(t), extend = TRUE)$surv[2]
VE.ITT.CI = 1-CI.V/CI.PB
VE.ITT.CI
```

The ITT effect using the hazard rate as risk measure can be estimated by fitting a Cox model:

```
data = data.jnj #Another dataset (e.g. data.pfizer) can also be used
#administrative censoring after day of estimating the VE
data$status = ifelse(data$time<t,yes=data$status,no=0)
#administrative censoring after day of estimating the VE
data$time = ifelse(data$time<t,yes=data$time,no=t)
#Cox proportional hazards model
fit = coxph(Surv(time, status) ~ X, data = data)
HR = exp(fit$coef["X"]) #Hazard ratio = exp(beta_X) in Cox model
```

```
VE.ITT.HR = 1-HR
VE.ITT.HR
```

The ITT effect using the incidence rate as risk measure can be estimated by fitting a Poisson model:

```
data = data.jnj #Another dataset (e.g. data.pfizer) can also be used
#administrative censoring after day of estimating the VE
data$status = ifelse(data$time<t,yes=data$status,no=0)
#administrative censoring after day of estimating the VE
data$time = ifelse(data$time<t,yes=data$time,no=t)
#logarithm of the follow-up time will be used as offset in the Poisson model
data$log_time = log(data$time)
#Poisson model
fit = glm(status~ X, offset=log_time,data=data,family=poisson)
IR = exp(fit$coef["X"]) #Incidence ratio = exp(beta_X) in Poisson model
VE.ITT.IR = 1-IR
VE.ITT.IR
```

##### D.2.2 PP effects (removing cases before $\alpha$)

In this section, we show how PP effects, where cases during the ramp-up time are removed from the analysis set, can be estimated.

We specify the day at which we will estimate the vaccine efficacy, for example at the end of the trial, and the length of the ramp-up period:

```
t = censor.day
#End of the ramp-up period (14 days in Janssen trial, 28 days in Pfizer trial)
alpha = 14
```

The data for the PP effects consist of all patients who were not infected during the ramp-up time:

```
data = data.jnj #Another dataset (e.g. data.pfizer) can also be used
data$PP.set = ifelse(data$time<alpha&data$status==1,yes=0,no=1) #indicator for PP set
data = subset(data,PP.set==1) #patients in dataset who were not infected in [0,alpha]
```

The PP effect using the cumulative incidence as risk measure can be estimated by fitting a Kaplan-Meier curve:

```
#Kaplan Meier survival curve
fit = survfit(Surv(time, status) ~ X, data = data)
#1-Survival probability in placebo arm of the PP dataset
CI.PB = 1-summary(fit,time=c(t), extend = TRUE)$surv[1]
```

```
#1-Survival probability in vaccine arm of the PP dataset
CI.V =  1-summary(fit,time=c(t), extend = TRUE)$surv[2]
VE.PPremove.CI = 1-CI.V/CI.PB
VE.PPremove.CI
```

The PP effect using the hazard rate as risk measure can be estimated by fitting a Cox model:

```
#administrative censoring after day of estimating the VE
data$status =ifelse(data$time<t,yes=data$status,no=0)
#administrative censoring after day of estimating the VE
data$time = ifelse(data$time<t,yes=data$time,no=t)
#Cox proportional hazards model
fit = coxph(Surv(time, status) ~ X, data = data)
HR = exp(fit$coef["X"]) #Hazard ratio = exp(beta_X) in Cox model
VE.PPremove.HR = 1-HR
VE.PPremove.HR
```

The PP effect using the incidence rate as risk measure can be estimated by fitting a Poisson model:

```
#administrative censoring after day of estimating the VE
data$status =ifelse(data$time<t,yes=data$status,no=0)
#administrative censoring after day of estimating the VE
data$time = ifelse(data$time<t,yes=data$time,no=t)
#logarithm of the follow-up time will be used as offset in the Poisson model
data$log_time = log(data$time)
#Poisson model
fit = glm(status~ X, offset=log_time,data=data,family=poisson)
IR = exp(fit$coef["X"]) #Incidence ratio = exp(beta_X) in Poisson model
VE.PPremove.IR = 1-IR
VE.PPremove.IR
```

##### D.2.3 PP effects (censoring cases before $\alpha$)

In this section, we show how PP effects, where cases during the ramp-up time are censored, can be estimated. We specify the day at which we will estimate the vaccine efficacy, for example at the end of the trial:

```
t = censor.day
#End of the ramp-up period (14 days in Janssen trial, 28 days in Pfizer trial)
alpha = 14
```

First, we censor all patients who were infected during the ramp-up time:

```
data = data.jnj #Another dataset (e.g. data.pfizer) can also be used
#administrative censoring after day of estimating the VE
data$status =ifelse(data$time<t,yes=data$status,no=0)
#administrative censoring after day of estimating the VE
data$time = ifelse(data$time<t,yes=data$time,no=t)
#censoring if infected before alpha
data$status =ifelse(data$time<alpha,yes=0,no=data$status)
```

The PP effect using the cumulative incidence as risk measure can be estimated by fitting a Kaplan-Meier curve:

```
fit = survfit(Surv(time, status) ~ X, data =data) #Kaplan Meier survival curve
#1-Survival probability in placebo arm of the PP dataset
CI.PB= 1-summary(fit,time=c(t), extend = TRUE)$surv[1]
#1-Survival probability in vaccine arm of the PP dataset
CI.V =  1-summary(fit,time=c(t), extend = TRUE)$surv[2]
VE.PPcensor.CI = 1-CI.V/CI.PB
VE.PPcensor.CI
```

The PP effect using the hazard rate as risk measure can be estimated by fitting a Cox model:

```
#Cox proportional hazards model
fit = coxph(Surv(time, status) ~ X, data = data)
HR = exp(fit$coef["X"]) #Hazard ratio = exp(beta_X) in Cox model
VE.PPcensor.HR = 1-HR
VE.PPcensor.HR
```

The PP effect using the incidence rate as risk measure can be estimated by fitting a Poisson model:

```
#logarithm of the follow-up time will be used as offset in the Poisson model
data$log_time = log(data$time)
#Poisson model
fit = glm(status~ X, offset=log_time,data=data,family=poisson)
IR = exp(fit$coef["X"]) #Incidence ratio = exp(beta_X) in Poisson model
VE.PPcensor.IR = 1-IR
VE.PPcensor.IR
```

##### D.2.4 VE if ramp-up period can be eliminated

Here, we show how the estimator for the hypothetical VE ‘if the ramp-up period can be eliminated’, as discussed in Appendix C.3.4, can be implemented in R.

**Method 1: Only estimating $\psi$**

We specify the dataset, the day at which we will estimate the vaccine efficacy, for example at the end of the trial, and the length of the ramp-up period:

```
data = data.jnj #Another dataset (e.g. data.pfizer) can also be used
t = censor.day
#End of the ramp-up period (14 days in Janssen trial, 28 days in Pfizer trial)
alpha = 14
```

1. First, we estimate a Kaplan-Meier curve:

   ```
   fit = survfit(Surv(time, status) ~ X, data = data)
   ```

2. Now we estimate $\psi$.

   (a) An initial estimate for $\psi$ is obtained by minimizing the squared difference in survival probabilities at the end of the trial.

   Therefore, we specify 3 functions. The first function returns the mapped survival probability under vaccination at a time $t$ using the SDM (equation (12)) for given parameters $\rho$, $\psi$ and $\alpha$:

   ```
   #fit: survival object fitted on the observed data
   #rho: specified parameter for the SDM model
   # (Indicates how much weaker the vaccine effect is during the ramp-up time.
   # Should be in the interval [0,1])
   #psi: specified parameter for the SDM model
   # (Represents the vaccine effect. Higher values mean higher efficacy.
   # 0 indicates no vaccine effect.)
   #alpha: specified parameter for the SDM model (Length of the ramp-up time)
   #t: time for which the mapped survival probability needs to be returned

   count.surv = function(fit,rho,psi,alpha,t){
   time1 = t/exp(rho*psi)
   surv1 = ifelse(t<alpha, yes=summary(fit,time=c(time1),
   extend = TRUE)$surv[1],no=0)
   time2 = (t+alpha*(exp(psi*(1-rho))-1))/exp(psi)
   surv2 = ifelse(t>=alpha,yes=summary(fit,time=c(time2),
    extend = TRUE)$surv[1],no=0)
   return(surv1+surv2)
   }
   ```

   The second function returns the squared difference between observed and mapped vaccine survival probability at time $t$. This function will be used to perform a grid search over a range of $\psi$ values ($\rho$ and $\alpha$ need to be specified):

```
#x: value for psi
#fit: survival object fitted on the observed data
#rho: specified parameter for the SDM model
# (Indicates how much weaker the vaccine effect is during the ramp-up time.
# Should be in the interval [0,1])
#alpha: specified parameter for the SDM model (Length of the ramp-up time)
#t: time for which the squared difference between observed vaccine
# and mapped vaccine survival probability needs to be returned

calculate.diff.1param = function(x,fit,rho,alpha,t){
psi = as.numeric(x[1])
#predict infection time under vaccine at time t using the SDM model
pred = count.surv(fit=fit,rho=rho,psi=psi,alpha=alpha,t=t)
#observed survival probability under vaccine at time t
obs = summary(fit,time=c(t), extend = TRUE)$surv[2]
#difference between the 2 probabilities
diff = pred-obs
output = diff^2
return(output)
}
```

The third function returns a start value for $\psi$ between 0 and `max.psi`. The mapped and observed survival functions are compared at time $t$ (using the squared difference). A grid search is performed for 100 $\psi$ values in the interval [0, `max.psi`] and the value for $\psi$ for which the difference between the mapped and observed survival function is minimized, is returned.

```
#data: observed dataset
#fit: survival object fitted on the observed data
#rho: specified parameter for the SDM model
# (Indicates how much weaker the vaccine effect is during the ramp-up time.
# Should be in the interval [0,1])
#alpha: specified parameter for the SDM model (Length of the ramp-up time)
#t: time for which the squared difference between observed and mapped
# vaccine survival probability needs to be calculated
#max.psi: maximum possible psi value

initial.1param = function(data,fit,rho,alpha,t,max.psi){
res <- gridSearch(fun=calculate.diff.1param,fit=fit,rho=rho,alpha=alpha,t=t,
lower = 0, upper = max.psi, npar = 1, n = 100)
initial.psi = res$minlevels
return(initial.psi)
```

```
}
```

Now we can estimate an initial value for $\psi$ by minimizing the squared difference in survival probabilities at the end of the trial. We specify a value for $\rho$ and $\alpha$ and a maximum value for $\psi$:

```
rho = 0
alpha = 14
max.psi = 10
initial.psi = initial.1param(data,fit,rho,alpha,t=t,max.psi=max.psi)
initial.psi
```

(b) This initial estimate is now updated by minimizing the squared difference in restricted mean survival times.

We specify again 3 functions. The first function simulates data under vaccine according to the SDM (equation (12)) with specified $\rho$, $\psi$ and $\alpha$ values. This simulated dataset will be used to calculate the restricted mean survival time.

```
#n: number of patients
#fit: survival object fitted on the observed data
#rho: specified parameter for the SDM model
# (Indicates how much weaker the vaccine effect is during the ramp-up time.
# Should be in the interval [0,1])
#psi: specified parameter for the SDM model
# (Represents the vaccine effect. Higher values mean higher efficacy.
# 0 indicates no vaccine effect.)
#alpha: specified parameter for the SDM model (Length of the ramp-up time)
#censor.day: last visit of the trial

counterfactual.data = function(n,fit,rho,psi,alpha,censor.day){
tdom<-seq(1,censor.day,by=1)
#failure times under vaccine
failtimes.V = c()
u = runif(n) #draws from uniform distribution
Surv =  c()  #survival function under vaccine
for(t in tdom){
time1 = t/exp(rho*psi)
surv1 = ifelse(t<alpha, yes=summary(fit,time=c(time1),
extend = TRUE)$surv[1],no=0)
time2 = (t+alpha*(exp(psi*(1-rho))-1))/exp(psi)
surv2 = ifelse(t>=alpha,yes=summary(fit,time=c(time2),
extend = TRUE)$surv[1],no=0)
Surv[t] = surv1+surv2
```

```
}
for(i in 1:n){
colsums = colSums(outer(Surv, u[i], '>'))
#Failure times are randomly drawn from the mapped survival function
failtimes.V[i] = ifelse(colsums==0, yes=0, no = tdom[colsums])
}
data= data.frame(patient=1:n,X=0,time=failtimes.V)
#Patients are administratively censored after last visit of the trial
data$status =ifelse(data$time<censor.day,yes=1,no=0)
return(data)
}
```

The second function returns squared difference in restricted mean survival time till time $L$. This function will be used to find a good estimate for $\psi$ and a penalty is added to make sure that the estimate is positive.

```
#x: value for psi
#fit: survival object fitted on the observed data
#data: observed dataset
#rho: specified parameter for the SDM model
# (Indicates how much weaker the vaccine effect is during the ramp-up time.
# Should be in the interval [0,1])
#alpha: specified parameter for the SDM model (Length of the ramp-up time)
#L: restricted mean survival time is estimated from baseline till time L

calculate.diff.RMST.1param = function(x,fit,data,rho,alpha,L){
psi = as.numeric(x[1])
 #simulated data under vaccine according to the SDM
data.count = counterfactual.data(n=20000,fit=fit,rho=rho,psi=psi,
alpha=alpha,censor.day=censor.day)
data.count$X = 0
data.long = rbind(subset(data,X==1),data.count)
#Restricted mean survival times for the observed and mapped vaccine arm
obj = rmst2(time=data.long$time,status=data.long$status,
arm=data.long$X,tau=L)
#Difference in restricted mean survival time
diff = obj$RMST.arm1$result["RMST","Est."]-obj$RMST.arm0$result["RMST","Est."]
output = diff^2 + ifelse(psi<0,yes=1000,no=0) #penalty if psi is negative
return(output)
}
```

The third function estimates a value for $\psi$ ($\rho$ and $\alpha$ need to be specified). Survival curves are compared using restricted mean survival time at time $L$.

```
#data: observed dataset
#fit: survival object fitted on the observed data
#initial.values = c(psi.initial) initial value for psi
#rho: specified parameter for the SDM model
# (Indicates how much weaker the vaccine effect is during the ramp-up time.
# Should be in the interval [0,1])
#alpha: specified parameter for the SDM model (Length of the ramp-up time)
#L: restricted mean survival time is estimated from baseline till time L

estimate.RMST.1param = function(data,fit,initial.values,rho,alpha,L){
psi = optim(par =initial.values,fn = calculate.diff.RMST.1param,fit=fit,
 data=data,rho=rho,alpha=alpha,L=L)$par
return(psi)
}
```

Now we can update the initial value for $\psi$ by minimizing the squared difference in in restricted mean survival times over the entire duration of the trial

```
psi = estimate.RMST.1param(data,fit,initial.values=c(initial.psi),
  rho,alpha,L=censor.day)
parameters = c(rho,psi,alpha)
names(parameters) = c("rho","psi","alpha")
parameters
```

To check how well the SDM with the obtained parameters approximates the observed survival curve, a Kaplan-Meier curve can be plotted. Therefore, we define a function that plots the original survival curves and the predicted vaccine curve for certain $\rho$, $\psi$ and $\alpha$.

```
#fit: survival object fitted on the observed data
#data: observed dataset
#rho: specified parameter for the SDM model
# (Indicates how much weaker the vaccine effect is during the ramp-up time.
# Should be in the interval [0,1])
#psi: specified parameter for the SDM model
# (Represents the vaccine effect. Higher values mean higher efficacy.
# 0 indicates no vaccine effect.)
#alpha: specified parameter for the SDM model (Length of the ramp-up time)
#censor.day: last visit of the trial

plot.SDM = function(fit,data,rho,psi,alpha,censor.day){
fit = survfit(Surv(time, status) ~ X, data = data)
data.count = counterfactual.data(n=100000,fit=fit,rho=rho,psi=psi,alpha=alpha,
censor.day=censor.day)
```

```
data.copy = data
data.copy$X = ifelse(data.copy$X==0,yes="Placebo",no="Vaccine")
data.count$X = "Modelled vaccine curve"
data.long <<- rbind(data.copy,data.count)
data.long$X <- factor(data.long$X,
levels=c("Placebo", "Vaccine", "Modelled vaccine curve"))
p = ggsurvplot(survfit(Surv(time, status) ~ X, data = data.long), title = " ",
xlab = "Days",
ylab = "Cumulative incidence",size=1,fun="event",
palette = c("red", "blue","pink"))$plot
p = p+ geom_vline(xintercept = alpha, linetype="dotted",  color = "red", size=1)
print(p)
}

rho = parameters["rho"]
psi = parameters["psi"]
alpha = parameters["alpha"]
plot.SDM(fit,data,rho,psi,alpha,censor.day)
```

Finally, the vaccine efficacy 'if the ramp-up time can be eliminated' can be calculated by setting the ramp-up period to 0 days:

```
rho = parameters["rho"]
psi = parameters["psi"]
#1-Survival probability in vaccine arm if there would be no ramp-up time (alpha=0)
CI.V = 1-count.surv(fit,rho,psi,alpha=0,t)
#1-Survival probability in placebo arm
CI.PB = 1-summary(fit,time=c(t),extend=TRUE)$surv[1]
VE.hypothetical.1param.CI = 1-(CI.V)/(CI.PB)
VE.hypothetical.1param.CI
```

**Method 2: Estimating $\psi$ and $\rho$**

We specify the dataset, the day at which we will estimate the vaccine efficacy, for example at the end of the trial, and the length of the ramp-up period:

```
data = data.jnj #Another dataset (e.g. data.pfizer) can also be used
t = censor.day
#End of the ramp-up period (14 days in Janssen trial, 28 days in Pfizer trial)
alpha = 14
```

1. First, we estimate a Kaplan-Meier curve:

   ```
   fit = survfit(Surv(time, status) ~ X, data = data)
   ```

2. Now we estimate $\rho$ and $\psi$.

   (a) Initial estimates for $\psi$ and $\rho$ are obtained by minimizing the sum of squared differences in survival probabilities at the end of the ramp-up period and the end of the trial.

   Therefore, we specify 2 functions. The first function returns the sum of the squared differences between observed and mapped vaccine survival probability at the visits in `t.vector`. This function will be used to perform a grid search over a range of $\psi$ and $\rho$ values ($\alpha$ needs to be specified):

```
#x: vector with values for rho and psi
#fit: survival object fitted on the observed data
#alpha: specified parameter for the SDM model (Length of the ramp-up time)
#t.vector: vector with times for which the squared difference between
# observed vaccine and mapped vaccine survival probability
# needs to be returned

calculate.diff.2param = function(x,fit,alpha,t.vector){
rho = as.numeric(x[1])
psi = as.numeric(x[2])
t1 = t.vector[1]
t2 = t.vector[2]
#predict infection time under vaccine at times t1 and t2 using the SDM model
pred1 = count.surv(fit=fit,rho=rho,psi=psi,alpha=alpha,t=t1)
pred2 = count.surv(fit=fit,rho=rho,psi=psi,alpha=alpha,t=t2)
#observed survival probabilities under vaccine at times t1 and t2
obs1 = summary(fit,time=c(t1), extend = TRUE)$surv[2]
obs2 = summary(fit,time=c(t2), extend = TRUE)$surv[2]
#difference between the 2 probabilities
diff1 = pred1-obs1
diff2 = pred2-obs2
output = diff1^2 + diff2^2 #sum of squared differences
return(output)
}
```

   The second function returns a start value for $\rho$ between 0 and `max.rho` and a start value for $\psi$ between 0 and `max.psi` ($\alpha$ needs to be specified). The mapped and observed survival functions are compared at the times in `t.vector` (using the sum of the squared differences). A grid search is performed for 50 $\rho$ values in the interval [0, `max.rho`] and 50 $\psi$ values in the interval [0, `max.psi`] and the values for $\rho$ and $\psi$ for which the sum of the squared differences between the mapped and observed survival function is minimized, are returned.

```
#data: observed dataset
#fit: survival object fitted on the observed data
#alpha: specified parameter for the SDM model (Length of the ramp-up time)
#t.vector: times for which the squared difference between observed
# and mapped vaccine survival probability needs to be calculated
#max.rho: maximum possible rho value
#max.psi: maximum possible psi value

initial.2param = function(data,fit,alpha,t.vector,max.rho,max.psi){
res <- gridSearch(fun=calculate.diff.2param,fit=fit,alpha=alpha,
t.vector=t.vector, lower = c(0,0), upper = c(max.rho,max.psi),
 npar = 2, n = 50)
initial.rho = res$minlevels[1]
initial.psi = res$minlevels[2]
output = c(initial.rho,initial.psi)
names(output) = c("rho","psi")
return(output)
}
```

Now we can estimate initial values for $\rho$ and $\psi$ by minimizing the sum of the squared differences in survival probabilities at the end of the ramp-up period and the end of the trial. We specify a value for $\alpha$ and a maximum value for $\rho$ and $\psi$:

```
max.rho = 1
max.psi = 10
t.vector = c(alpha,censor.day)
initial = initial.2param(data, fit,alpha,t.vector=t.vector,
max.rho=max.rho,max.psi=max.psi)
initial.rho = initial["rho"]
initial.rho
initial.psi = initial["psi"]
initial.psi
```

(b) These initial estimates are now updated by minimizing the sum of the squared difference in restricted mean survival times at the end and halfway through the study.

We specify again 2 functions. The first function returns the sum of the squared differences in restricted mean survival time till the first and second time in `L.vector`. This function will be used to find good estimates for $\rho$ and $\psi$ and a penalty is added to make sure that the estimate of $\psi$ is positive and the estimate of $\rho$ in the interval $[0, 1]$.

```
#x: vector with values for rho and psi
#fit: survival object fitted on the observed data
#data: observed dataset
#alpha: specified parameter for the SDM model (Length of the ramp-up time)
#L.vector: vector with two visits, restricted mean survival time is
# estimated from baseline till time these visits

calculate.diff.RMST.2param = function(x,fit,data,alpha,L.vector){
rho = as.numeric(x[1])
psi = as.numeric(x[2])
#simulated data under vaccine according to the SDM
data.count = counterfactual.data(n=20000,fit=fit,rho=rho,psi=psi,
alpha=alpha,censor.day=censor.day)
data.count$X = 0
data.long = rbind(subset(data,X==1),data.count)
#Restricted mean survival times for the observed and mapped vaccine arm
# from baseline till first visit in L.vector
obj = rmst2(time=data.long$time,status=data.long$status,
arm=data.long$X,tau=L.vector[1])
#Difference in restricted mean survival time
diff1 = obj$RMST.arm1$result["RMST","Est."]
-obj$RMST.arm0$result["RMST","Est."]
#Restricted mean survival times for the observed and mapped vaccine arm
# from baseline till second visit in L.vector
obj = rmst2(time=data.long$time,status=data.long$status,
arm=data.long$X,tau=L.vector[2])
#Difference in restricted mean survival time
diff2 = obj$RMST.arm1$result["RMST","Est."]
-obj$RMST.arm0$result["RMST","Est."]
#sum of squared differences
#and penalties if psi is negative or rho outside [0,1]
output =diff1^2 + diff2^2 +ifelse(psi<0,yes=1000,no=0)
+ifelse(rho<0,yes=1000,no=0)+ifelse(rho>1,yes=1000,no=0)
return(output)
}
```

The second function estimates values for $\rho$ and $\psi$ ($\alpha$ needs to be specified). Survival curves are compared using restricted mean survival time at the first and second time in `L.vector`.

```
#data: observed dataset
#fit: survival object fitted on the observed data
#initial.values: vector with initial values for rho and psi
#alpha: specified parameter for the SDM model (Length of the ramp-up time)
#L.vector: vector with two visits, restricted mean survival time
# is estimated from baseline till these visits

estimate.RMST.2param = function(data,fit,initial.values,alpha,L.vector){
param = optim(par =initial.values,fn = calculate.diff.RMST.2param
,fit=fit,data=data,alpha=alpha,L=L.vector)$par
param = c(param,alpha)
names(param) = c("rho","psi","alpha")
return(param)
}
```

Now we can update the initial values for $\rho$ and $\psi$ by minimizing the sum of the squared differences in restricted mean survival times over half and the entire duration of the trial

```
L.vector = c(censor.day/2,censor.day)
param =  estimate.RMST.2param(data,fit,initial.values
=c(initial.rho,initial.psi),alpha = alpha,L.vector=L.vector)
param = c(param)
names(param) = c("rho","psi","alpha")
```

To check how well the SDM with the obtained parameters approximates the observed survival curve, a Kaplan-Meier curve can be plotted.

```
rho = parameters["rho"]
psi = parameters["psi"]
alpha = parameters["alpha"]
plot.SDM(fit,data,rho,psi,alpha,censor.day)
```

Finally, the vaccine efficacy ‘if the ramp-up time can be eliminated’ can be calculated by setting the ramp-up period to 0 days:

```
#1-Survival probability in vaccine arm if there would be no ramp-up time (alpha=0)
CI.V = 1-count.surv(fit,rho,psi,alpha=0,t)
#1-Survival probability in placebo arm
CI.PB = 1-summary(fit,time=c(t),extend=TRUE)$surv[1]
VE.hypothetical.2param.CI = 1-(CI.V)/(CI.PB)
VE.hypothetical.2param.CI
```

**Method 3: Estimating $\psi$, $\rho$ and $\alpha$**

We specify the dataset, the day at which we will estimate the vaccine efficacy, for example at the end of the trial, and the length of the ramp-up period:

```
data = data.jnj #Another dataset (e.g. data.pfizer) can also be used
t = censor.day
#End of the ramp-up period (14 days in Janssen trial, 28 days in Pfizer trial)
alpha = 14
```

1. First, we estimate a Kaplan-Meier curve:

   ```
   fit = survfit(Surv(time, status) ~ X, data = data)
   ```

2. Now we estimate $\psi$, $\rho$ and $\alpha$.

   (a) Initial estimates for $\psi$, $\rho$ and $\alpha$ are obtained by minimizing the sum of squared differences in survival probabilities at a quarter of the length of the study, halfway through the study and at the end of the study.

   Therefore, we specify 2 functions. The first function returns the sum of the squared differences between observed and mapped vaccine survival probability at the visits in `t.vector`. This function will be used to perform a grid search over a range of $\psi$, $\rho$ and $\alpha$ values:

   ```
   #x: vector with values for rho, psi and alpha
   #fit: survival object fitted on the observed data
   #t.vector: vector with times for which the squared difference between
   # observed vaccine and mapped vaccine survival probability
   # needs to be returned

   calculate.diff.3param = function(x,fit,t.vector){
   rho = as.numeric(x[1])
   psi = as.numeric(x[2])
   alpha = as.numeric(x[3])
   t1 = t.vector[1]
   t2 = t.vector[2]
   t3 = t.vector[3]
   #predict infection time under vaccine at times t1, t2 and t3
   #using the SDM model
   pred1 = count.surv(fit=fit,rho=rho,psi=psi,alpha=alpha,t=t1)
   pred2 = count.surv(fit=fit,rho=rho,psi=psi,alpha=alpha,t=t2)
   pred3 = count.surv(fit=fit,rho=rho,psi=psi,alpha=alpha,t=t3)
   #observed survival probabilities under vaccine at times t1, t2 and t3
   ```

```
obs1 = summary(fit,time=c(t1), extend = TRUE)$surv[2]
obs2 = summary(fit,time=c(t2), extend = TRUE)$surv[2]
obs3 = summary(fit,time=c(t3), extend = TRUE)$surv[2]
#difference between the 2 probabilities
diff1 = pred1-obs1
diff2 = pred2-obs2
diff3 = pred3-obs3
output = diff1^2 + diff2^2 +diff3^2  #sum of squared differences
return(output)
}
```

The second function returns a start value for $\rho$ between 0 and `max.rho`, a start value for $\psi$ between 0 and `max.psi` and a start value for $\alpha$ between 0 and `max.alpha`. The mapped and observed survival functions are compared at the times in `t.vector` (using the sum of the squared differences). A grid search is performed for 20 $\rho$ values in the interval [0, `max.rho`], 20 $\psi$ values in the interval [0, `max.psi`] and 20 $\alpha$ values in the interval [0, `max.alpha`] and the values for $\rho$, $\psi$ and $\alpha$ for which the sum of the squared differences between the mapped and observed survival function is minimized, are returned.

```
#data: observed dataset
#fit: survival object fitted on the observed data
#t.vector: vector with times for which the squared difference
# between observed and mapped vaccine survival probability needs
# to be calculated
#max.rho: maximum possible rho value
#max.psi: maximum possible psi value
#max.alpha: maximum possible alpha value

initial.3param = function(data,fit,t.vector,max.rho,max.psi,max.alpha){
res <- gridSearch(fun=calculate.diff.3param,fit=fit,t.vector=t.vector,
 lower = c(0,0,0), upper = c(max.rho,max.psi,max.alpha), npar = 3, n = 20)
initial.rho = res$minlevels[1]
initial.psi = res$minlevels[2]
initial.alpha = res$minlevels[3]
output = c(initial.rho,initial.psi,initial.alpha)
names(output) = c("rho","psi","alpha")
return(output)
}
```

Now we can estimate initial values for $\rho$, $\psi$ and $\alpha$ by minimizing the sum of the squared differences in survival probabilities at a quarter of the length of the study, halfway through the study and at the end of the study. We specify a maximum value for $\rho$, $\psi$ and $\alpha$:

```
max.rho = 1
max.psi = 10
max.alpha = censor.day/4
t.vector = c(censor.day/4,censor.day/2,censor.day)
initial = initial.3param(data, fit,t.vector=t.vector,
 max.rho=max.rho,max.psi=max.psi,max.alpha = max.alpha)
initial.rho = initial["rho"]
initial.rho
initial.psi = initial["psi"]
initial.psi
initial.alpha = initial["alpha"]
initial.alpha
```

(b) These initial estimates are now updated by minimizing the sum of the squared difference in restricted mean survival times at a quarter of the length of the study, halfway through the study and at the end of the study.

We specify again 2 functions. The first function returns the sum of the squared differences in restricted mean survival time till the first, second and third time in `L.vector`. This function will be used to find good estimates for $\rho$, $\psi$ and $\alpha$ and a penalty is added to make sure that the estimate of $\psi$ is positive, the estimate of $\rho$ in the interval $[0, 1]$ and the estimate of $\alpha$ not larger than half of the study length.

```
#x: vector with values for rho, psi and alpha
#fit: survival object fitted on the observed data
#data: observed dataset
#L.vector: vector with three visits, restricted mean survival time
# is estimated from baseline till  these visits

calculate.diff.RMST.3param = function(x,fit,data,L.vector){
rho = as.numeric(x[1])
psi = as.numeric(x[2])
alpha = as.numeric(x[3])
#simulated data under vaccine according to the SDM
data.count = counterfactual.data(n=20000,fit=fit,rho=rho,psi=psi,
alpha=alpha,censor.day=censor.day)
data.count$X = 0
data.long = rbind(subset(data,X==1),data.count)
#Restricted mean survival times for the observed and mapped
#vaccine arm from baseline till first visit in L.vector
```

```
obj = rmst2(time=data.long$time,status=data.long$status,
arm=data.long$X,tau=L.vector[1])
#Difference in restricted mean survival time
diff1 = obj$RMST.arm1$result["RMST","Est."]
-obj$RMST.arm0$result["RMST","Est."]
#Restricted mean survival times for the observed and mapped
#vaccine arm from baseline till second visit in L.vector
obj = rmst2(time=data.long$time,status=data.long$status,
arm=data.long$X,tau=L.vector[2])
#Difference in restricted mean survival time
diff2 = obj$RMST.arm1$result["RMST","Est."]
-obj$RMST.arm0$result["RMST","Est."]
#Restricted mean survival times for the observed and mapped
#vaccine arm from baseline till third visit in L.vector
obj = rmst2(time=data.long$time,status=data.long$status,
arm=data.long$X,tau=L.vector[3])
#Difference in restricted mean survival time
diff3 = obj$RMST.arm1$result["RMST","Est."]
-obj$RMST.arm0$result["RMST","Est."]
#sum of squared differences and penalties if psi is negative
#or rho outside [0,1] or alpha outside [0,censor.day/2]
output =diff1^2 + diff2^2 + diff3^2 +ifelse(psi<0,yes=1000,no=0)
 +ifelse(rho<0,yes=1000,no=0)+ifelse(rho>1,yes=1000,no=0)
 +ifelse(alpha<0,yes=1000,no=0)+ifelse(alpha>censor.day/2,yes=1000,no=0)
return(output)
}
```

The second function estimates values for $\rho$, $\psi$ and $\alpha$. Survival curves are compared using restricted mean survival time at the first and second time in `L.vector`.

```
#data: observed dataset
#fit: survival object fitted on the observed data
#initial.values: vector with initial values for rho, psi and alpha
#L.vector: vector with three visits, restricted mean survival time
# is estimated from baseline till these visits

estimate.RMST.3param = function(data,fit,initial.values,L.vector){
param = optim(par =initial.values,fn = calculate.diff.RMST.3param,
fit=fit,data=data,L=L.vector)$par
param = c(param)
names(param) = c("rho","psi","alpha")
```

```
return(param)
}
```

Now we can update the initial values for $\rho$, $\psi$ and $\alpha$ by minimizing the sum of the squared differences in restricted mean survival times a quarter of the length of the study, halfway through the study and at the end of the study.

```
L.vector = c(censor.day/4,censor.day/2,censor.day)
param = estimate.RMST.3param(data,fit,initial.values
=c(initial.rho,initial.psi,initial.alpha),L.vector=L.vector)
param = c(param)
names(param) = c("rho","psi","alpha")
```

To check how well the SDM with the obtained parameters approximates the observed survival curve, a Kaplan-Meier curve can be plotted.

```
rho = parameters["rho"]
psi = parameters["psi"]
alpha = parameters["alpha"]
plot.SDM(fit,data,rho,psi,alpha,censor.day)
```

Finally, the vaccine efficacy 'if the ramp-up time can be eliminated' can be calculated by setting the ramp-up period to 0 days:

```
#1-Survival probability in vaccine arm if there would be no ramp-up time (alpha=0)
CI.V = 1-count.surv(fit,rho,psi,alpha=0,t)
#1-Survival probability in placebo arm
CI.PB = 1-summary(fit,time=c(t),extend=TRUE)$surv[1]
VE.hypothetical.3param.CI = 1-(CI.V)/(CI.PB)
VE.hypothetical.3param.CI
```
